# Multi-omics analysis in primary T cells elucidates mechanisms behind disease associated genetic loci

**DOI:** 10.1101/2023.07.19.23292550

**Authors:** Chenfu Shi, Danyun Zhao, Stefano Rossi, Antonios Frantzeskos, James Ding, Carlo Ferrazzano, Charlotte Wynn, Ryan Hum, Ellie Richards, Muskan Gupta, Chuan Fu Yap, Darren Plant, Richard Grencis, Paul Martin, Antony Adamson, Stephen Eyre, John Bowes, Anne Barton, Pauline Ho, Magnus Rattray, Gisela Orozco

## Abstract

In this study, we present the most extensive dataset of chromatin conformation data with matching gene expression and chromatin accessibility from primary T cells to date. We use this data to enhance our understanding of the different mechanisms by which GWAS variants impact gene regulation and revealing how natural genetic variation alter chromatin accessibility and structure in primary cells at an unprecedented scale. Capitalizing on this vast dataset, we refine the mapping of GWAS loci to implicated regulatory elements, such as CTCF binding sites and other enhancer elements, aiding gene assignment. Importantly, we uncover *BCL2L11* as the probable causal gene within the RA locus rs13396472, despite the GWAS variants’ intronic positioning relative to *ACOXL* and we identify mechanisms involving *SESN3* dysregulation in the RA locus rs4409785. Given these genes’ significant role in T cell development and maturation, our work is vital for deepening our comprehension of autoimmune disease pathogenesis and suggesting potential treatment targets.

## INTRODUCTION

Genome wide association studies (GWAS) have uncovered a large proportion of the genetic basis underlying most common traits and diseases (Buniello et al. 2019), such as psoriatic arthritis (PsA) (Soomro et al. 2022). It is widely established that these genetic associations are implicated mainly in the regulation of genes rather than altering coding sequences directly (Farh et al. 2015; Finucane et al. 2015). Because regulatory elements perturbed by these variants can affect genes located far away from their genomic location with complex, cell-type specific, mechanisms (Nolis et al. 2009; Shlyueva et al. 2014; Kundaje et al. 2015; Simeonov et al. 2017; Alasoo et al. 2018), understanding the functional role of these variants involves significant effort and currently remains the main challenge in the field (Cano-Gamez and Trynka 2020; Orozco et al. 2022).

Many studies have used functional genomics techniques to study risk loci, both to identify the causal variants among the ones in strong linkage disequilibrium (LD), and to assign candidate target genes (Orozco 2022). These include using epigenomic markers to identify the variants that are likely to affect regulatory elements in disease relevant cell types (Kundaje et al. 2015; Robertson et al. 2021), eQTL methods (Schmiedel et al. 2018; Võsa et al. 2021), quantifying co-activation of enhancers and promoters (Delaneau et al. 2019; Boix et al. 2021) and using chromatin conformation capture methods to assign enhancers to genes (Martin et al. 2015; Mifsud et al. 2015; Cairns et al. 2016; McGovern et al. 2016; Mumbach et al. 2017; Choy et al. 2018; Montefiori et al. 2018; Greenwald et al. 2019; Miguel-Escalada et al. 2019; Ray-Jones et al. 2020; Shi et al. 2021; Ge et al. 2021). Most of these studies have, however, used cell lines or healthy lineages due to the difficulty of generating Hi-C libraries from primary cells and tissues, but there is extensive evidence that gene regulation and chromatin conformation is highly cell state specific (Schmitt et al. 2016; Yang et al. 2020; Iqbal et al. 2021). These studies also typically used small samples sizes due to the cost of these techniques, and the only report examining the effect of natural DNA variation on 3D chromatin structure used cell lines and a technique with low resolution (Gorkin et al. 2019). Finally, many of these studies used capture techniques to reduce cost, such as ChIA-PET (Fullwood et al. 2010), HiChIP (Mumbach et al. 2017) or capture Hi-C (Mifsud et al. 2015), which limit the view of the local chromatin structure.

Here we address these issues by generating the largest collection of high quality, high resolution, Hi-C maps in primary T cells using the commercial Arima Hi-C protocol, which allowed for higher quality and reproducibility whilst reducing the input material required, enabling the use of primary cells isolated from patients. We chose PsA as an exemplar disease as this autoimmune condition has a strong, complex genetic component and high heritability (O’Rielly and Rahman 2011; Veale and Fearon 2018). Additionally, the mechanisms underlying this disease are not very well understood and have not been the focus of previous functional genomics studies.

We interpret the results from our study using the latest understanding of how chromatin conformation is linked to gene regulation, validating recently published gene regulatory mechanisms (Rowley and Corces 2018; Hsieh et al. 2020; Krietenstein et al. 2020; Hua et al. 2021; Aljahani et al. 2022; Goel et al. 2023). We study how chromatin conformation varies across a population of individuals in different cell types and conditions and with genotype, at a significantly higher resolution to what has been previously achieved (Gorkin et al. 2019). Finally, we show how this rich dataset allows us to decipher the mechanisms by which disease associated variants increase the risk of developing disease by studying a selection of GWAS loci from a variety of immune conditions.

## RESULTS

### A compendium of functional genomics data in primary T cells

In this study, we present the largest dataset to date of matching chromatin conformation, chromatin accessibility and gene expression from freshly isolated primary CD4^+^ and CD8^+^ T cells from 55 PsA patients and 19 Healthy controls (Figure 1A). This consists of 108 Hi-C libraries (49 billion reads), 128 RNA-seq libraries and 126 ATAC-seq libraries. Uniform Manifold Approximation and Projection (UMAP) plots show that the samples separate by cell type, with the synovial fluid samples forming a separate cluster from the peripheral blood samples (Figure 1B-E). We do not detect a clear separation between samples derived from PsA patients and healthy controls (Figure 1B-E), but we see a slight separation by sex of the individuals (Supplementary Figure 1).

**Figure 1.**
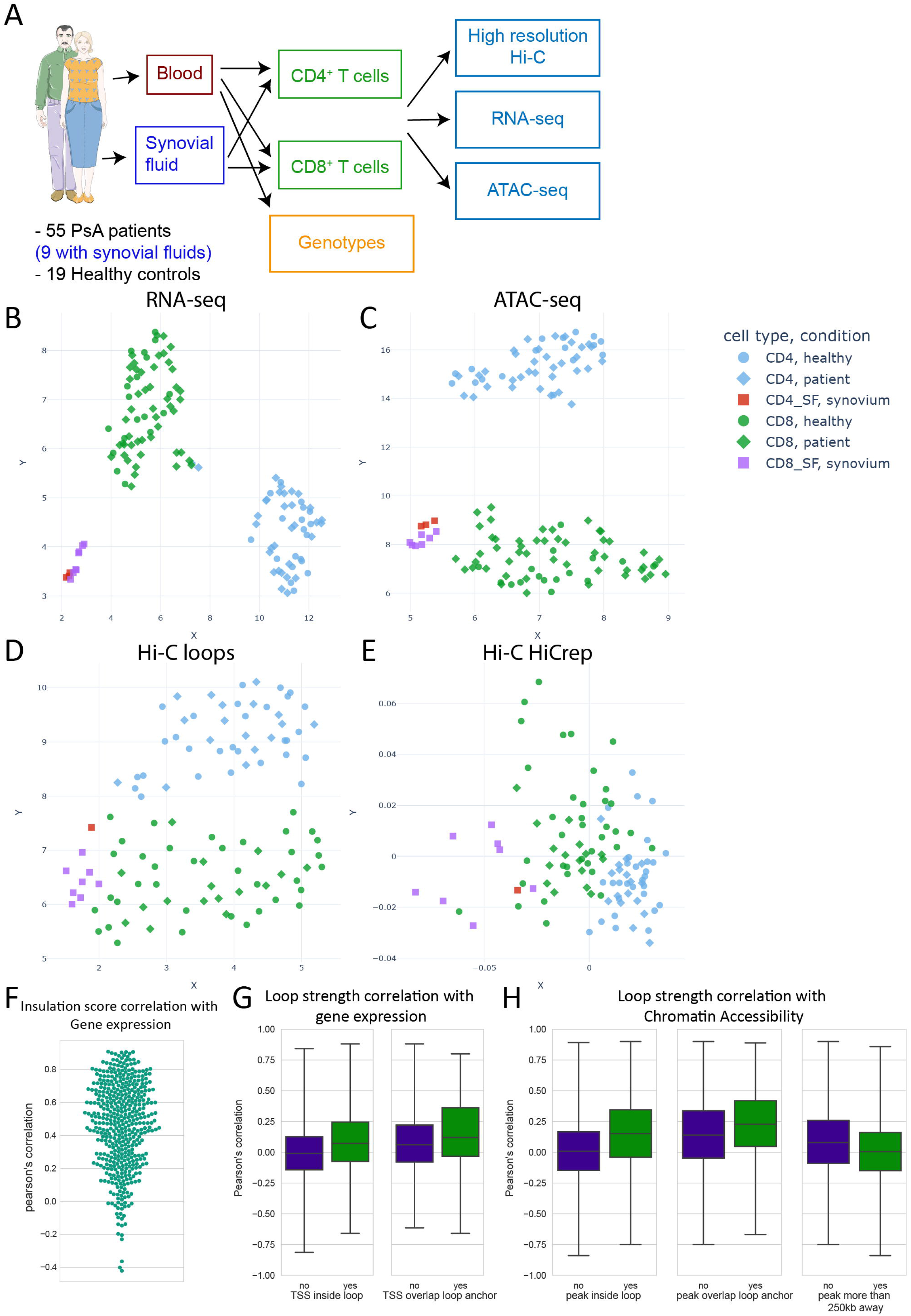
A) Summary of the study design. B) UMAP of RNA-seq data, counts from DESeq2 C) UMAP of the ATAC-seq data, counts from DiffBind. D) UMAP of the Hi-C loops data. E) Multidimensional Scaling plot (MDS) of HiCRep analysis of the Hi-C data. F) Pearson’s correlation between gene expression and insulation score of the bin overlapping promoter of the gene. Each dot is a gene. G) Pearson’s correlation between gene expression and loop strength. Left: comparison for loops that surround the gene vs loops that are nearby the gene. Right: comparison between loops that surround the gene with loops for which loop anchor overlaps the promoter of the gene. H) Pearson’s correlation between height of ATAC-seq peaks and loop strength. Left: Comparison between loops that surround the peak vs loops that are nearby the peak. Middle: Comparison between loops that surround the peak with loops for which loop anchor overlaps the peak. Right: Comparison between loops that are nearby the peak and loops that are more than 250kb away.

We identify chromatin loops using a recently developed loop caller specifically designed for high resolution contact maps, Mustache (Ardakany et al. 2020). We identify 69,303, 72,884, 30,415 and 12,000 loops from merged maps from CD4^+^, CD8^+^, CD8^+^ synovial fluid and CD4^+^ synovial fluid T cells respectively, which are then used for further analysis. As expected, 75% of these loops overlap a CTCF peak in at least one anchor, with stronger loops more likely to overlap a CTCF peak, although only 42% overlap a CTCF peak at both anchors (Supplementary Figure 2A-C). Additionally, we find that 89% of loops overlap a chromatin accessibility peak at one end, 75% of loops overlap one at both ends and 47% of loops overlap a transcription start site (TSS).

Although it is known that immortalized cell lines differ significantly from primary cells, it has never been shown how altered is the chromatin conformation. Here, we generated matching Hi-C libraries from two cell lines, Jurkat CD4^+^ T cells and MyLa CD8^+^ T cells to compare them to our primary T cells datasets. Previous studies have shown that topologically associating domains (TADs) are generally stable between different cell types and differentiation states (Dixon et al. 2015). In line with these findings, we find that around 95% of TAD boundaries from MyLa and Jurkat cells are present in their corresponding primary cells and 99% of TAD boundaries from primary cells are present in the cell lines. For reference, two primary T cell samples share around 97% of TAD boundaries. However, we can identify large differences when looking at the compactness of these TADs. We estimate, using an outlier detection technique (see methods), that 45% of the genome has significant differences in insulation score (Supplementary Figure 2D). We also see large differences when we analyse high resolution loops. We find that up to 20% of all loops are differentially interacting in cell lines compared to primary cells (Supplementary Figure 2E). These regions with altered chromatin conformation include genes that are important in T-cell function such as the *ANKRD44, KLRF1, ITK* and *GZMB* genes (Supplementary Figure 3-4) and regions that overlap disease associated GWAS loci such as the PsA loci rs11743851 (*SLC22A5*) or rs13203885 (*FYN*) (Supplementary Figure 5**Error! Reference source not found.**).

### Chromatin conformation is highly associated with gene regulation

Next, we explored how chromatin conformation varies across the cell populations studied. Out of 105,956 loops, 29,454 were differentially interacting between CD8^+^ and CD4^+^ T cells (FDR<0.1), and out of 113,350 insulation score bins at 25kb, 41,614 bins have differential compactness. Out of the 86,720 combined loops, 15,597 were differentially interacting between CD8^+^ T cells isolated from blood and synovial fluid (FDR<0.1), together with 30,408 bins with differential compactness.

Although we could identify 166 (CD4^+^) and 38 (CD8^+^) genes differentially expressed between patients with active disease and remission and 110 (CD4^+^) and 437 (CD8^+^) genes between active disease and healthy (Table 1), we could not identify any significant differential looping or differential insulation between these subgroups. This is likely because the differences between these subgroups are significantly smaller compared to the differences between cell types (Table 1), and the techniques used did not allow for the identification of these subtle changes between heterogenous groups.

**Table 1.** Number of differentially expressed genes and differentially bound ATAC-seq peaks.

| Comparison | Differentially expressed genes | Differentially bound ATAC-seq peaks |
| --- | --- | --- |
| Active vs non-active disease CD8<sup>+</sup> T cells | 38 | 3 |
| Active vs non-active disease CD4<sup>+</sup> T cells | 166 | 1463 |
| Active disease vs controls CD8<sup>+</sup> T cells | 437 | 127 |
| Active disease vs controls CD4<sup>+</sup> T cells | 110 | 116 |
| CD8<sup>+</sup> vs CD4<sup>+</sup> T cells | 10109 | 43648 |
| CD8<sup>+</sup> SF vs CD8<sup>+</sup> Active disease Blood | 12709 | 30079 |
| CD4<sup>+</sup> SF vs CD4<sup>+</sup> Active disease Blood | 11536 | 37411 |

We next probed the interplay between gene expression and chromatin conformation. Our data shows a strong positive correlation between the insulation score of the chromatin domain and gene expression (Figure 1F). Expanding on this finding, we examined the correlation between gene expression and the strength of the chromatin loops. As expected, we observed a strong positive correlation when a loop overlapped the gene promoter on one of the anchors. Interestingly, loops that encompassed a gene promoter, but not at the loop anchors, also showed correlation, albeit to a lesser extent (Figure 1G). This indicates that indirect spatial relationships between promoters and loop anchors are associated with gene expression. Next, we included chromatin accessibility in the analysis. In this context, ATAC-seq peaks within a loop but that did not directly overlap the loop anchors exhibited a significant positive correlation with the loops, surprisingly even stronger than that observed for TSS (Figure 1H). This emphasizes the possible regulatory role of accessible chromatin regions, irrespective of their spatial relation to loop anchors, further extending our understanding of the chromatin conformation’s influence on gene expression. These findings collectively suggest a nuanced role for chromatin architecture in transcriptional regulation.

Delving deeper into specific regions, our data enables us to visualize intricate local shifts in chromatin conformation. For instance, within the *IL7R* gene region (Figure 2A), which is located in a large TAD which contains *SPEF2* and *CAPSL* genes as well, we observed complex changes associated with increased interactions around the subTAD encompassing *IL7R*. This increase in compactness is associated with a decrease of interactions outside of this subTAD, but an increase of interactions upstream to an intron of *SPEF2*. We find that the chromatin accessibility peaks located in this region are correlated with the expression of *IL7R*, suggesting a complex orchestration of chromatin interactions which regulate gene expression. We find that these complex changes in local chromatin conformation are very common across many important genes and our data can help understand the mechanisms involved in the regulation of these genes.

**Figure 2.**
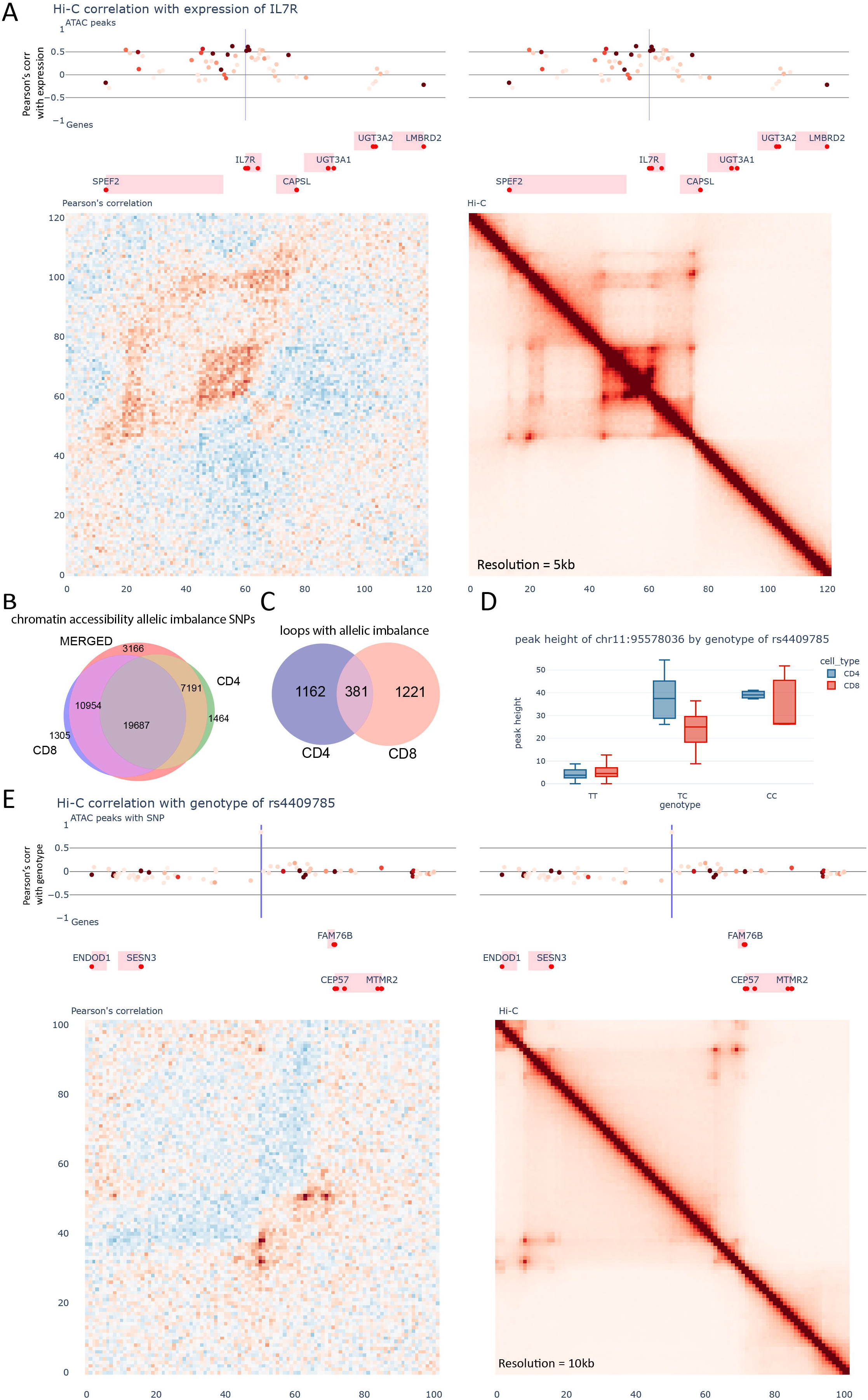
A) Visualization of the region surrounding *IL7R*. Top: ATAC-seq peaks. Intensity of the colour indicates the average peak-height of the peak, whilst position across the Y axis indicates the pearson’s correlation with the expression of *IL7R*. Middle: Genes from Gencode v29. The red dots indicate the transcription start sites. Bottom left: Correlation between the Hi-C contacts and the expression of *IL7R*. Bottom right: Merged Hi-C map. B) Venn diagram showcasing the overlap between chromatin accessibility allelic imbalance results from CD4^+^ T cells, CD8^+^ T cells and merged dataset. C) Venn diagram showcasing the overlap between loop allelic imbalance results from CD4^+^ T cells and CD8^+^ T cells. D) Accessibility of CTCF site located at chr11:95578036 vs the genotype of rs4409785. E) Visualization of the region surrounding rs4409785. Top: ATAC-seq peaks. Intensity of the colour indicates the average peak-height of the peak, whilst position across the Y axis indicates the pearson’s correlation with the genotype of rs4409785. Middle: Genes from Gencode v29. The red dots indicate the transcription start sites. Bottom left: Correlation between the Hi-C contacts and the genotype of rs4409785. Bottom right: Merged Hi-C map.

### Natural genetic variation is strongly associated with alterations in gene expression, chromatin accessibility and chromatin conformation

Building upon these findings, we proceeded to assess the impact of common genetic variants on gene expression, chromatin accessibility, and chromatin conformation. To this aim, we implemented insulation score QTL (insQTL) and loop strength QTL (loopQTL) in the context of chromatin conformation, to investigate the influence on the strength and insulation of topologically associating domains (TADs) and the strength of chromatin loops respectively. In conjunction, chromatin accessibility QTL (caQTL) was utilized to discern how genetic variation affects the functionality of enhancers and other regulatory elements and eQTL for gene expression. The number of significant loops, insulation score bins, chromatin accessibility peaks, and genes with a significant QTL are presented in table 2.

**Table 2.**
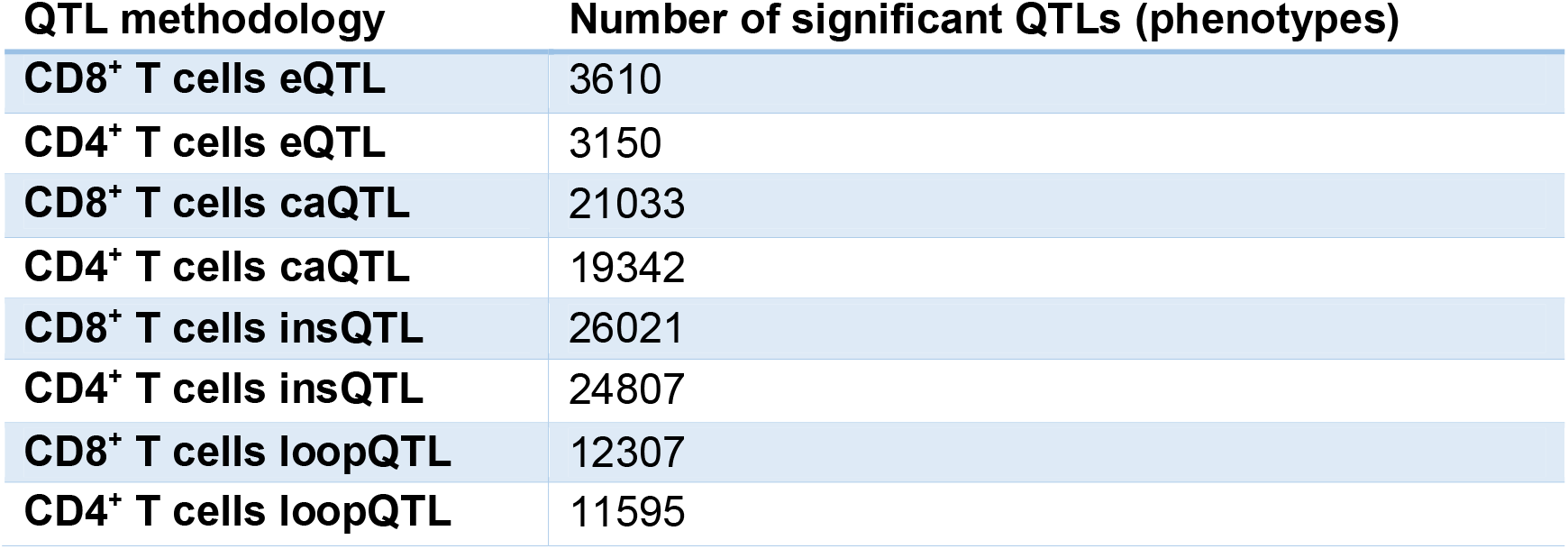
Number of significant QTLs for each of the methodologies tested.

Next, we explored how these different QTL results intersect with one another. Initially, we compared the results from the same modality across the two cell types (CD8^+^ and CD4^+^ T cells). For eQTLs and caQTLs we found an overlap of 27.2% and 26.6% respectively, with 100% concordance in effect directionality for shared QTLs. In the case of insQTLs, a smaller portion of significant QTLs (∼19%) were shared, while for loopQTLs, the overlap further diminished to 6%, with a maintained concordance at 100%. This suggests that loopQTL discovery may be more challenging than eQTLs and caQTLs due to subtler changes in chromatin conformation. Additionally, these results indicate that, whilst our study is the largest of its kind for chromatin conformation and is among the largest for chromatin accessibility QTLs, the sensitivity remains significantly limited.

Continuing our investigation, we looked into how QTLs from different modalities overlapped each other. We could recover approximately 10% of eQTLs from the caQTLs that overlapped the promoters of the eQTL genes, with a concordance rate of about 91%. Similar levels of overlap and concordance were found between eQTLs and insQTLs and caQTLs and insQTLs. These findings are in line with the results of our previous section, corroborating the notion that stronger gene expression is associated with increased chromatin domain insulation.

When we considered the overlap between eQTLs and caQTLs with loopQTLs, the concordance dropped to 67% and 78% respectively, if we included all loops for which the promoter or the ATAC-peak was located between the two loop anchors. However, the concordance increased, reaching 85% and 90% respectively, when we included only the loops that overlapped the promoter or the ATAC-peak at loop anchors. This suggests that loops that directly overlap the promoter of the genes or the ATAC-peak are more likely to be positively correlated with the expression of the genes and the activity of the peak. Meanwhile, loops that don’t overlap the promoter or the ATAC-peak, whilst still significant QTLs, don’t necessarily have a positive correlation, indicating different mechanisms at play.

An alternative method to study the effect of a genetic variant with a molecular phenotype is allelic imbalance. This approach calculates the proportion of signal derived from each allele in heterozygous individuals and can also be helpful in pinpointing the causal variants within the ones in LD. Because a large proportion of GWAS SNPs are predicted to affect enhancers, we apply this technique to chromatin accessibility regions to identify enhancers altered by these variants. We identify 40,997 variants with allelic imbalance in chromatin accessibility (FDR<0.1) out of 243,664 variants overlapping open chromatin regions in the merged dataset (Figure 2B). We find that allelic imbalance produces slightly different results although we find that it is generally more sensitive than QTL analysis (see methods for more information). We could recover 38% of caQTL that directly overlap their feature, but caQTL could only recover 13% of significant allelic imbalance variants. For the variants that overlap between the two analyses, we find 98% concordance in their directionality. Additionally, we have applied this technique to our chromatin conformation data, identifying 2,764 loops with allelic imbalance across the two cell types (Figure 2C). In this case we find that only 5% of the loopQTLs have their associated variant within their loop anchors and the majority of the loops don’t have enough sequencing depth to call allelic imbalance significantly (albeit with 100% concordance in directionality for the significant ones). These results suggest that although the QTL and allelic imbalance strongly agree on the effect, they can be used to interrogate a different set of mechanisms and can be combined to increase our understanding of GWAS loci.

### Unveiling the consequence of GWAS variants that affect the binding of CTCF at an unprecedent resolution

One of the most important proteins in the regulation of chromatin architecture is CTCF. For this reason, we searched for variants within CTCF binding sites in the allelic imbalance loops and loopQTLs. We find that 15% of loops with allelic imbalance have one variant located within a CTCF peak. We also find that 7% of loopQTLs and 10% of insQTL have at least one variant overlapping a CTCF peak. These results indicate that even though a majority of these structures are defined by CTCF related mechanisms, the way genetic variants affect these interactions is not primarily driven by alteration of the CTCF binding. Finally, we find that about 20% of GWAS loci have one variant overlapping a CTCF peak.

For example, in the RA locus tagged by rs4409785, we find that the lead SNP is a strong caQTL for the peak it maps to (FDR 5.52e-07) (Figure 2D), as well as being in allelic imbalance (FDR 1.01e-03). Previous studies have revealed that the risk allele (ALT) of this SNP creates a new CTCF binding site (Sadowski et al. 2019). Using our data we can visualize, for the first time, how the creation of this new CTCF binding site alters the local chromatin structure (Figure 2E). This SNP is located at the centre of a large chromatin domain, which contains the gene *SESN3* and a large gene desert. The risk allele of rs4409785 is associated with the creation of two new loops, not visible in the merged high resolution Hi-C, which connect this element to the downstream boundary of the chromatin domain. This results in an increase of the contacts within this new subTAD, and a reduction of interactions between this region and the upstream region which includes the *SESN3* gene. In this way, this reduces the ability of regulatory elements positioned downstream of rs4409785 to activate the gene *SESN3*. Public eQTL data shows that the risk allele of rs4409785 is associated with a reduction of expression of *SESN3* (Võsa et al. 2021). This risk locus had been previously assigned to the *CEP57* gene (Ishigaki et al. 2022). However, *SESN3* could have an important role in arthritis. A recent study suggests that SESN3^+^ memory T cells, a subset not extensively studied before, may play a significant role in the development of arthritis or autoimmune diseases. It was found that these cells, which were identified in skin biopsies from patients with psoriasis, could differentiate into cytotoxic CD4^+^ T cells, potentially contributing to the pathogenesis of autoimmune diseases such as rheumatoid arthritis (Argyriou et al. 2022).

Another interesting example is the *IKZF3*/*GSDMB*/*ORMDL3* GWAS locus, which is linked to many autoimmune diseases, including RA, Asthma, Systemic Sclerosis (SSc) and others (Ferreira et al. 2019; López-Isac et al. 2019; Ishigaki et al. 2022). Previous studies have localized the causal SNP as rs12936231 (Verlaan et al. 2009; Schmiedel et al. 2016; Sadowski et al. 2019), which creates a novel CTCF binding motif. We find in our dataset that this SNP is a strong caQTL for the ATAC-peak (p-value 5.02e-09 for CD4 and 2.38e-10 for CD8) (Figure 3A) and displays strong loop allelic imbalance (FDR 4.48e-25 for CD4 and 3.55e-33 for CD8) and multiple loopQTLs (one example plotted in Figure 3B). Although it is known that this novel CTCF binding site causes a change in chromatin looping, using our dataset, we can visualize the effect on this locus at an unprecedented detail. We find significant changes in the insulation of two contact domains (subTADs), associated with a reduction of interactions between them, whilst significantly increasing the interactions that go downstream of the new CTCF site (Figure 3C). Our allelic imbalance data also shows that other variants upstream of rs12936231, and in LD with it, have increased chromatin accessibility which indicates that this reduction of interactions results in an increase in activity of these enhancers. Together with eQTL data, which indicate upregulation of genes upstream of the CTCF motif (*IKZF3*) and downregulation of genes downstream (*GSDMB*, *ORMDL3*), we can conclude that this variant increases disease risk by disrupting the ability of upstream regulatory elements to contact these downstream genes.

**Figure 3.**
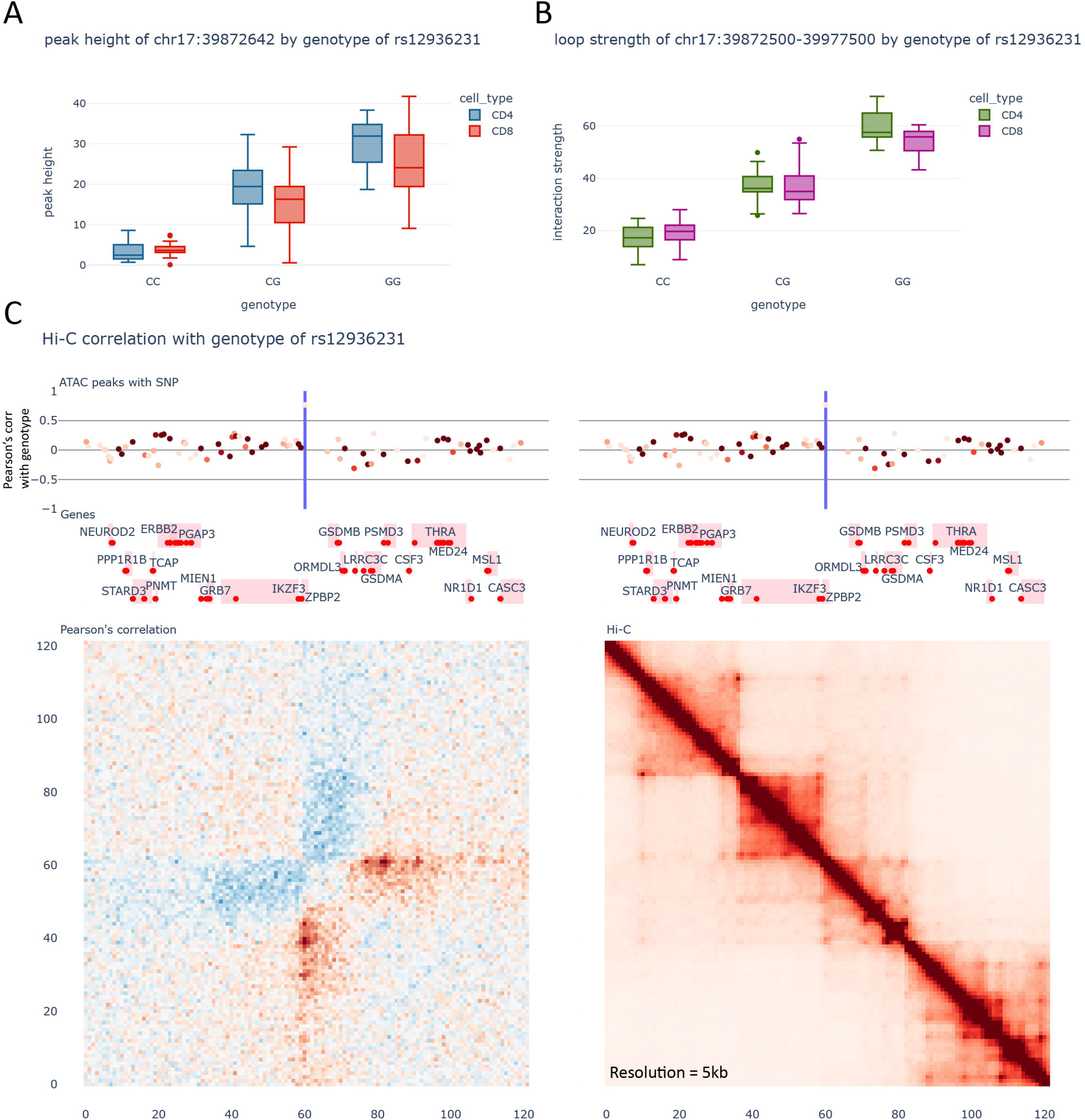
A) Accessibility of CTCF site located at chr17:39872642 vs the genotype of rs12936231. B) Representative loopQTL with the genotype of rs12936231 in the region. Loop displayed is chr17:39872500-39977500. C) Visualization of the region surrounding rs12936231. Top: ATAC-seq peaks. Intensity of the colour indicates the average peak-height of the peak, whilst position across the Y axis indicates the pearson’s correlation with the genotype of rs12936231. Middle: Genes from Gencode v29. The red dots indicate the transcription start sites. Bottom left: Correlation between the Hi-C contacts and the genotype of rs12936231. Bottom right: Merged Hi-C map.

Another region with a similar mechanism is a locus associated with white blood cell count and neutrophil counts indexed by the SNP rs10424217. We find that the SNP in LD rs2353678, directly overlaps a predicted CTCF binding motif, with our data showing a strong increase in chromatin accessibility both through allelic imbalance (p-value 2.99e-16) and caQTL (FDR 2.37e-175) (Supplementary Figure 6A). Similarly, to the previous region, this new CTCF site increases the separation between two chromatin domains (Supplementary Figure 6B). In this case, this is associated with an increased expression of most genes in this region, including *RPS16* (upstream), *SUPT5H*, *EID2*, *EID2B*, *TIMM50* (downstream), but a decrease of expression of *PLEKHG2* (upstream).

In summary, we show how some variants can affect the regulation of multiple genes, by affecting key regulatory elements that define chromatin architecture at an unprecedent detail and resolution. However, the majority of the GWAS loci do not act through this mechanism. We explore how our data can help us investigate GWAS loci with other mechanisms in the following section.

### Uncovering the mechanisms by which GWAS loci affect gene regulation by altering enhancer function

Previous studies have indicated that the majority of GWAS loci affect enhancers elements (Farh et al. 2015). Motivated by these findings, we utilized our dataset to investigate the mechanisms involved behind this class of loci by first using chromatin accessibility allelic imbalance as a tool to functionally fine-map GWAS loci (supplementary tables). Our analysis led to the identification of one or more allelic imbalance SNPs in about a third of the loci. For instance, in the *KLF13* psoriasis locus linked by rs28624578, out of the 4 SNPs in LD, 3 of them overlap chromatin accessibility regions. However, our data shows that only rs28510484 exhibits strong allelic imbalance (FDR 9.11E-36) with a 2.2 fold increase in chromatin accessibility. These SNPs are located in an intron of *KLF13* and eQTLgen data (Võsa et al. 2021) show an increase in *KLF13* expression associated with the risk (ALT) allele of rs28510484. Another example is the *RAB2A-CHD7* SSc locus linked by rs685985, where the SNPs are located between *RAB2A* and *CHD7*. Here, although 5 of the 7 SNPs in LD overlap open chromatin regions, only two have significant allelic imbalance, with one of the two having significantly stronger allelic imbalance (rs612558 FDR 3.98E-40, rs686852 FDR 5.4E-3). Interestingly, although the risk (ALT) allele of rs612558 is associated with a 25% reduction in chromatin accessibility, eQTLgen data indicate increased expression of *RAB2A*.

Remarkably, certain regions also display alterations in chromatin conformation. The RA locus linked by SNP rs7943728 serves as an example, wherein the SNP in LD rs968567 is the only SNP that overlaps chromatin accessibility regions and showcases pronounced allelic imbalance accompanied by an 8.2-fold increase in chromatin accessibility (FDR 0) for the protective (ALT) allele. This SNP is a strong caQTL for 4 separate ATAC-peaks, indicating that the alternate form of rs968567 causes an increased activity of other regulatory elements in the locus as well (Figure 4A-D). This strong increase in the activity of these elements results in the increase in interactions in the local chromatin architecture, particularly those bringing the genes *FADS2*, *FADS1*, *FADS3* and *FTH1* in closer contact with the enhancer elements affected by rs968567 (Figure 4E). Moreover, this variant is correlated with an increase in the expression of *FADS2*, *FADS1* (Figure 4F-G). These genes are crucial for the pathogenesis of RA as they play a key role in the biosynthesis of long-chain polyunsaturated fatty acids, like omega-3 and omega-6 (Hastings et al. 2001), known to influence inflammation and immune response mechanisms pivotal in the disease’s pathogenesis (Kostoglou-Athanassiou et al. 2020).

**Figure 4.**
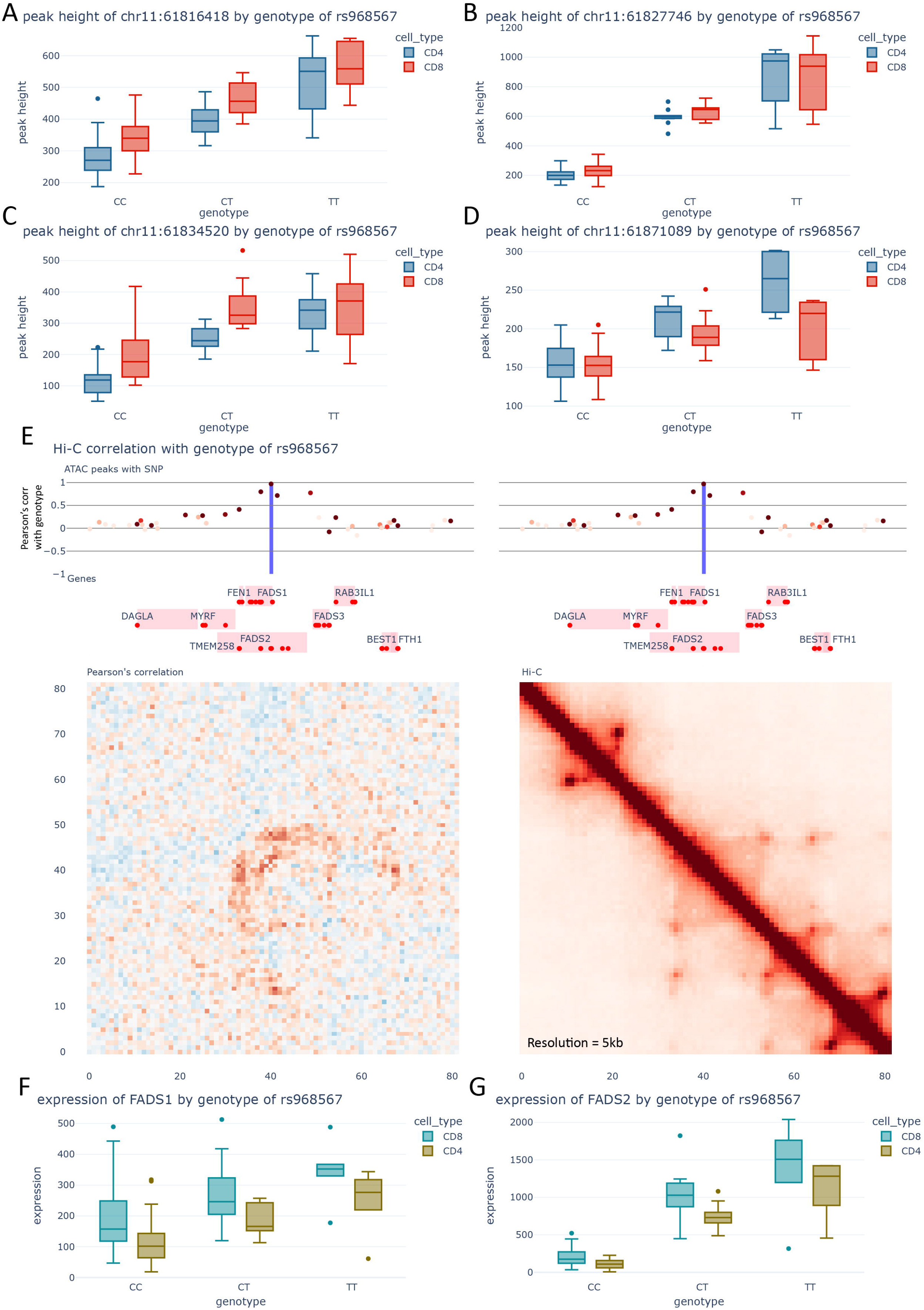
A) Chromatin accessibility of peak chr11:61816418 vs the genotype of rs968567. B) Chromatin accessibility of peak chr11:61827746 vs the genotype of rs968567. C) Chromatin accessibility of peak chr11:61834520 vs the genotype of rs968567.D) Chromatin accessibility of peak chr11:61871089 vs the genotype of rs968567. E) Visualization of the region surrounding rs968567. Top: ATAC-seq peaks. Intensity of the colour indicates the average peak-height of the peak, whilst position across the Y axis indicates the pearson’s correlation with the genotype of rs968567. Middle: Genes from Gencode v29. The red dots indicate the transcription start sites. Bottom left: Correlation between the Hi-C contacts and the genotype of rs968567. Bottom right: Merged Hi-C map. F) Expression of *FADS1* with the genotype of rs968567. G) Expression of *FADS2* with the genotype of rs968567.

Lastly, in the RA locus linked by rs13396472, only one SNP, rs13401811, shows strong allelic imbalance (FDR 1.86E-96), with a reduction of 40% in chromatin accessibility associated with the protective (ALT) allele. This SNP is also a strong caQTL for the overlapping ATAC-seq peak (p-value 1.43e-06, Figure 5A). This locus has been previous linked to *ACOXL* because the SNP in LD rs1554005, is a missense variant for *ACOXL*. Additionally, rs13401811 is located in an intron of *ACOXL*. However, the function of this gene is not especially intriguing for RA pathogenesis and is not expressed in T cells (less than 5 reads in our data) or other immune cells according to the DICE database (Schmiedel et al. 2018). Our chromatin conformation data reveals a loop connecting the enhancer affected by rs13401811 to the promoter of *BCL2L11* a gene located more than 300kb downstream of the GWAS SNPs. In fact, the activity of this enhancer appears to be highly correlated with the strength of the loop (p-value 1.25E-15, R2 0.47, Figure 5B-C).

**Figure 5.**
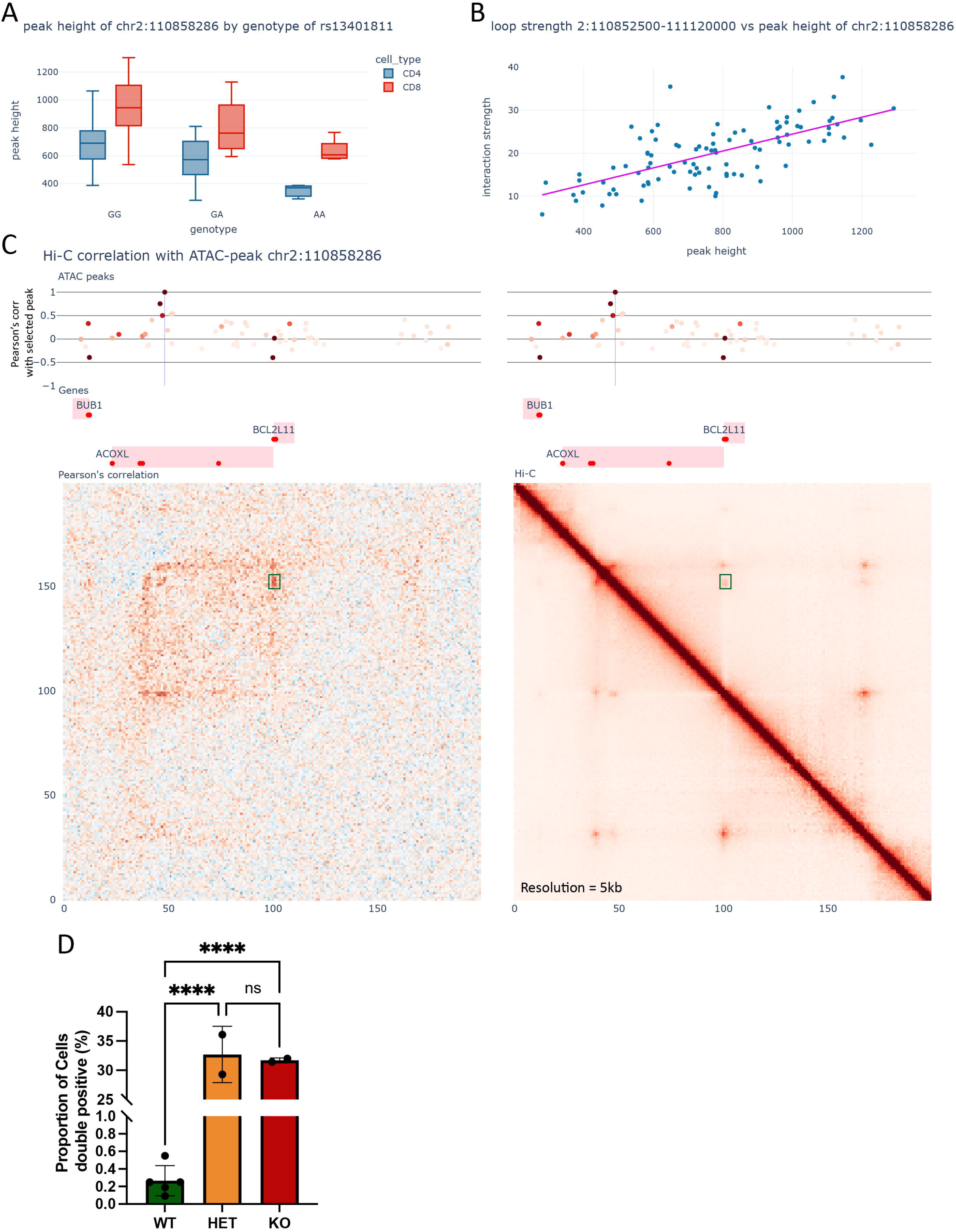
A) Chromatin accessibility of peak chr2:110858286 vs the genotype of rs13401811. B) Loop strength of the loop connecting the enhancer with the promoter of *BCL2L11* vs the peak height of the enhancer. C) Visualization of the region surrounding rs13401811. Top: ATAC-seq peaks. Intensity of the colour indicates the average peak-height of the peak, whilst position across the Y axis indicates the pearson’s correlation with the peak-height of the enhancer chr2:110858286. Middle: Genes from Gencode v29. The red dots indicate the transcription start sites. Bottom left: Correlation between the Hi-C contacts and the peak height of the enhancer chr2:110858286. Bottom right: Merged Hi-C map. The green box indicates the loop that connects the enhancer with the promoter of BCL2L11. D) Proportion of double-positive T cells from mouse spleen with deleted enhancer region.

*BCL2L11* has a critical role within the immune system, acting as a pro-apoptotic stimulator and modulating thymic negative selection. Knockout mice for *BCL2L11* display progressive autoimmune disease (Bouillet et al. 1999). To further study the in-vivo effects of the altered enhancer activity we decided to create a deletion of this regulatory region (hg38: chr2:110843317-110860033) located in the *ACOXL* intron in mouse (mm39: chr2: 127755856-127771494). Region comparison using Ensembl illustrates that the whole region, containing the genes *BUB1*, *ACOXL* and *BCL2L11*, as well as the regulatory elements deleted, are highly conserved between humans and mouse (Supplementary Figure 7). Preliminary results show that in mice of approximately 7.5 months of age there is a marked increase in the presence of a population of CD4/CD8 double-positive splenic T cells in both heterozygous and knockout mice when compared with their wild-type equivalents (Figure 5D). This indicates a possible evasion of central tolerance mechanisms in T lymphocytes. A smaller region overlapping our targeted regulatory locus has been previously knocked out in mice (Hojo et al. 2019). showing that this enhancer specifically alters the expression of *BCL2L11* in TCR-dependent activation of T cells, further strengthening our finding that rs13401811 increases the risk of developing RA by altering activation of *BCL2L11*.

## DISCUSSION

In this study, we have produced the most extensive dataset to date combining chromatin conformation, chromatin accessibility, and gene expression data from primary T cells isolated from Psoriatic Arthritis (PsA) patients and healthy individuals. Our data allowed us to clearly distinguish T cell samples into the different populations. However, we found that chromatin conformation variations between patients and controls were subtle and hard to detect, in line with previous attempts in the field (González-Serna et al. 2023).

Our work corroborates the recent findings about gene regulatory mechanisms and the relationship between chromatin conformation and gene regulation (Rowley and Corces 2018; Hsieh et al. 2020; Krietenstein et al. 2020; Hua et al. 2021; Aljahani et al. 2022; Goel et al. 2023). In that regard, we observed a strong correlation between chromatin compactness, as measured by insulation score, and gene expression, alongside looping strength. Additionally, our dataset enabled us to discern subtle alterations in local chromatin conformation linked to genotype, gene expression, and chromatin accessibility peaks, thus revealing intricate dynamics that remain hidden in conventional merged data.

We also present the most extensive analysis to date on the impact of genetic variation on chromatin conformation at an unprecedent resolution. By using complementary results from chromatin accessibility and gene expression we illustrate how variants that increase gene expression and chromatin accessibility also increase chromatin compactness and have intricate relationships with chromatin loops. Our results, however, indicate that our sample size is not sufficiently large to detect all QTLs, highlighting the ongoing need for larger-scale studies in this domain.

Finally, we leveraged our data to functionally refine GWAS loci. The interpretation of risk variants from GWAS has often been a stumbling block in their application to drug development. Compared to other studies, ours provides the most comprehensive functional context around these variants and underlines the complex nature of the mechanisms by which GWAS loci can influence gene regulation. The majority of prior studies relied on simple chromatin loops or raw contact frequency to associate genes with enhancers (Martin et al. 2015; Mifsud et al. 2015; Javierre et al. 2016; McGovern et al. 2016; Mumbach et al. 2017; Choy et al. 2018; Montefiori et al. 2018; Greenwald et al. 2019; Miguel-Escalada et al. 2019; Ray-Jones et al. 2020; Ge et al. 2021; Shi et al. 2021). However, this approach poses several challenges. Firstly, the majority of chromatin loops observed in Hi-C maps are structural loops mediated by CTCF, which do not link promoters with enhancers. Moreover, the majority of chromatin conformation methods are bulk methods, which means that enhancer-promoter interactions that appear only in a subpopulation of the cells, or that are more transient (van Staalduinen et al. 2023), might be significantly harder to detect. In that regard, our correlation method allows for the identification of dynamic structures and changes in interaction frequency that are not visible in the merged Hi-C map, such as those depicted in the *FADS1/2* locus. Other techniques, for example those based on Micro-C (Krietenstein et al. 2020; Aljahani et al. 2022; Goel et al. 2023) have lower background noise compared to Hi-C but are significantly more challenging technically, requiring large number of cells, and, to our knowledge, no study to date has been published that uses primary cells. Moreover, difficulties in the reproducibility of the libraries limit the kind of correlation and QTL analysis that have been presented here. Secondly, focusing on enhancer promoter interactions overlooks the exploration of other mechanisms by which genetic variants impact gene regulation and chromatin conformation, such as the CTCF binding altering variants presented in our study. These regions represent just the beginning of understanding the intricate mechanisms at play, such as the ability of specific variants to affect multiple genes at once and for regulatory elements to affect genes across TAD boundaries. This highlights the need to move beyond strict, static definitions of TADs, loops, and correlations in assigning genes to GWAS loci. Additionally, we identified notable differences between primary cells and cell lines, which suggest that caution needs to be taken when using cell lines to draw conclusions.

The mechanisms and genes we identified here through our dataset, such as *BCL2L11* and *SESN3*, present promising targets for novel, more effective therapeutics, and aid in patient stratification. New treatments that have genetic support are twice as likely to be approved (King et al. 2019). *BCL2L11*, is a key player in the apoptosis pathway and fundamental for T cell maturation (Bouillet et al. 1999; Hojo et al. 2019). Methotrexate, a widely used drug in rheumatology, has an inhibitory effect on dihydrofolate reductase (*DHFR*) which can influence various pathways, including those involving apoptosis where *BCL2L11* plays a significant role. In this context, our findings about *BCL2L11* regulation or expression could provide novel avenues for future research, including the exploration of how variations in *BCL2L11* regulation might affect therapeutic outcomes or the development of new treatments that target *BCL2L11* directly. Another breakthrough from our study is the identification of the potential mechanism by which the RA locus rs4409785 affects the expression of *SESN3*, a gene that plays a pivotal role in a specific subset of T cells implicated in the pathogenesis of autoimmune diseases (Argyriou et al. 2022). This discovery suggests the potential for novel therapies that target these cells directly.

In conclusion, our research elucidates the complex dynamics of chromatin conformation and gene regulation, offering potential implications for disease understanding and therapeutic development. We trust that our dataset will be a valuable resource for researchers in this field. To facilitate further research, we have made available all pre-processed data, including precomputed chromatin conformation maps, correlations with gene expression, chromatin accessibility, genotype, QTL datasets and code to replicate our analysis and annotate further GWAS results at http://bartzabel.ls.manchester.ac.uk/orozcolab/SNP2Mechanism/

## ONLINE METHODS

### Sample collection and cell isolation

Between 40ml and 100ml of peripheral blood was collected following informed consent from 55 PsA patients and 19 healthy controls and processed on the same day. Patients and healthy controls were recruited under the National Repository study ethics approval number IRAS 32231, MREC 99/8/084. PBMCs were isolated using Ficoll density gradient centrifugation. EasySep Human CD4^+^ and CD8^+^ T cell negative selection kits (StemCell technologies #17952 and #17953) were used to isolate CD4^+^ and CD8^+^ T cells respectively. Cells were then aliquoted and prepared for long-term storage separately for each technique used.

Synovial fluid extracted from patients with PsA was first treated with hyaluronidase enzyme (Sigma Aldrich #H3884) and then processed in a similar way as the peripheral blood samples.

To confirm that there was no sample mislabelling we checked the concordance of the genotype with the bam file by using the QTLtools v1.3.1 mbv function (Fort et al. 2017). To identify mismatches between CD4^+^ T cells and CD8^+^ T cells we used the gene expression of the *CD4* and *CD8A* genes for the RNA-seq data, the chromatin accessibility of the promoters of *CD4* and *CD8A* for ATAC-seq, and the loop strength of the loops surrounding the *CD4* gene for the Hi-C data. Sample mismatches were then corrected in the metadata files.

### Hi-C library preparation and sequencing

Approximately 3-5 million cells were crosslinked in 2% formaldehyde for 10 minutes at room temperature and the reaction was then quenched using a solution glycine + BSA. Samples were then washed in PBS and snap frozen on dry ice and then transferred to −80°C for long-term storage.

2 million cells were then used for library preparation following the two-restriction enzyme Arima Hi-C kit (Arima Genomics) and the KAPA HyperPrep kit (Roche #KK8504) manufacturer’s protocol using Illumina TruSeq dual indexes.

Library size was checked by TapeStation 4200 (Agilent) and QC was done by quantstudio (Life technologies) using the NEBNext® Library Quant Kit for Illumina (E7630). Sequencing was performed on the NovaSeq6000 platform using NovaSeq S4 flow cells at a target depth of 350 million reads per library and generating 150bp paired-end reads.

### RNA-seq library preparation and sequencing

Approximately 500 thousand to 1 million cells were lyzed and stored in 700uL Qiazol lysis reagent (QIAGEN, ref: 79306). To isolate RNA, 140 uL of chloroform was added. After centrifugation at 12000 x g for 15 min, approximately 350 uL of the upper layer containing the RNA was transferred and mixed with 350 uL of 70% ethanol. RNA isolation was continued from this point using the RNeasy MinElute kit (QIAGEN, ref: 74204) protocol. RNA libraries were then generated by NovoGene UK using a custom protocol based on cDNA synthesis using random hexamer primers and polyA mRNA enrichment. Sequencing was performed on the NovaSeq6000 platform using NovaSeq S4 flow cells at a target depth of 40 million reads per library and generating 150bp paired-end reads.

### ATAC-seq library preparation and sequencing

Approximately 500 thousand cells were frozen in Recovery™ Cell Culture Freezing Medium (Thermofisher #12648010) and stored at −80°C. An aliquot of 50 thousand cells was then used for ATAC-seq library preparation following the Omni-ATAC protocol as released by the Kaersten lab (Corces et al. 2017; Amanda Ackermann 2019).

Library size was checked by TapeStation 4200 (Agilent) and QC was done by quantstudio (Life technologies) using the NEBNext® Library Quant Kit for Illumina (E7630). Sequencing was performed on the NextSeq2000 platform at a target depth of 50 million reads per library and generating 50bp paired-end reads.

### Hi-C data processing, loop calling and clustering

Hi-C reads were quality filtered and trimmed and adapters were removed using fastp v0.20.1 (Chen et al. 2018). Reads were then processed and mapped to the GRCh38 genome using HiC-pro v3.0.0 (Servant et al. 2015), using default settings. Hic juicebox files for visualization and analysis were generated using the hicpro2juicebox.sh script using juicer tools v1.22.01. Cool files were generated from the juicebox files using hic2cool (https://github.com/4dn-dcic/hic2cool). TAD calling was done using onTAD (An et al. 2019) with default settings and ICE normalized 40kb resolution maps. Visualizations were made using a modified version of the python package coolbox (Xu et al. 2021) or through custom scripts available at (#####). The Hi-C processing pipeline is available on GitHub (https://github.com/ChenfuShi/hic_master_pipeline).

Loop calling was performed on merged libraries by cell population (CD4^+^, CD8^+^ and CD8^+^ synovial fluid) using Mustache (Ardakany et al. 2020) at a resolution of 2.5kb and SCALE normalization. This tool allows the detection of locally enriched loops (similar to HICCUPS (Rao et al. 2014)) rather than defining a model of contact decay based on genomic distance (such as FitHiC or CHiCAGO (Cairns et al. 2016; Bhattacharyya et al. 2019)) but it has shown to have improved sensitivity, although the number of loops identified strongly correlates with the depth of the contact maps. A common list of loops was obtained by merging the loops lists and removing duplicates. Counts for each loop from each sample were extracted at 5kb resolution and including the surrounding pixels, in such a way that counts were extracted from a 15kb-by-15kb region around the centre of the loop. These counts were extracted from SCALE normalized maps and library normalization between samples was computed by extracting the expected cis counts based on genomic distance decay and multiplying the counts by the ratio between the sample’s decay distribution with a reference sample. This distance decay function was calculated using cooltools v0.5.1 (Venev et al. 2022) api function “expected_cis” at a 10kb resolution and calculated for each chromosome. Except these normalization steps, no batch effect corrections were found to be necessary or influenced the results significantly.

Clustering of Hi-C libraries was performed using two separate methods. The first method is based on the the loops called. Normalized counts were simply treated as data matrix and standard scaled, after which a UMAP transformation (McInnes et al. 2018) was applied to the matrix to obtain the 2 component representation of the data. Alternatively, a PCA transformation could also be applied obtaining similar results. The second method was based on HiCrep (Yang et al. 2017). We used a fast python implementation (Lin et al. 2021) which allowed the computation of pairwise distances across all samples. Distances were calculated using a bin size of 50kb, a smoothing factor h of 5, and a maximum genomic distance of 5mb. We then applied MDS scaling on the pairwise distance matrix to generate the 2D plot in the results. Results are visualized for chromosome 1.

### RNA-seq data processing and analysis

RNA-seq reads were quality filtered and trimmed and adapters were removed using fastp v0.20.1 (Chen et al. 2018). Counts per transcript were generated using salmon v1.6.0 (Patro et al. 2017), with a decoy aware transcriptome (Gencode hg38 v29) and the validate mappings option enabled. Counts were then aggregated per gene and differential analysis was performed using DESeq2 (Love et al. 2014). Genes with average counts lower than 5 were removed. In all cases, unless otherwise specified, sex was used as a covariate. For other analysis, normalized counts were extracted from DESeq2 and used. No batch effect correction was found to be necessary.

### ATAC-seq data processing and analysis

ATAC-seq reads were quality filtered and trimmed and adapters were removed using fastp v0.20.1 (Chen et al. 2018). Reads were then mapped using bowtie2 v2.4.4 (Langmead and Salzberg 2012) using the very-sensitive pre-set. Duplicates were removed using the MarkDuplicates functionality of picard tools (Institute 2019) and mitochondrial reads were removed, as well as removing low quality alignments (MAPQ<30) and keeping only properly paired reads. Coverages were generated from cleaned alignments using the bamCoverage functionality from DeepTools2 (Ramírez et al. 2016). Peaks were called using macs2 v2.2.7.1 (Zhang et al. 2008). The ATAC-seq processing pipeline is available on GitHub (https://github.com/ChenfuShi/ATAC_ChIP_pipeline).

Data for all samples was then analysed in R using DiffBind (Stark and Brown 2011). A consensus peakset was generated using peaks that were present in at least 5 samples. Peak size was set to 500bp. Cross library normalization was performed using the DESeq2 normalization method (DBA_NORM_NATIVE) but using only reads in peaks (DBA_LIBSIZE_PEAKREADS), as different samples had significantly different proportion of reads in peaks, which would bias the default normalization method. Counts were extracted at this point for further analysis, except for differential binding analysis, which was performed within DiffBind, using the DESeq2 method. No batch effect correction was found to be necessary. We found that quantile normalization slightly improved results, such as separation of cell types on the PCA plot and correlation between expression and peak height of promoter peaks, and as such we used quantile normalized counts for all downstream analysis.

We have also used the ATAC-seq data to call genotypes for the samples for which we did not have Hi-C data. This was done using a similar method as the Hi-C data using GLIMPSE v1.1.1 (Rubinacci et al. 2021).

### Hi-C loop differential analysis

We developed a novel method based on linear regression to perform differential analysis from a large dataset of Hi-C data. All previous tools failed to compute the large number of samples used in our experiment. We extracted and normalized counts as described in the previous sections using a distributed system across our computational cluster. For each loop we then applied an ordinary least square regression using the python package statsmodels (Seabold and Perktold 2010). In the comparisons presented here the gender of the individual was set as a co-variate. Adjusted p-values were calculated using FDR correction (Benjamin-Hochberg) as implemented in the statsmodels package. A permutation run of the data (shuffling the cell type labels) resulted in less than 3 significant differential loops.

### Differential insulation score analysis

We calculated insulation score using the cooltools v0.5.1 (Venev et al. 2022) api function “insulation” using SCALE normalized matrixes at 25kb resolution and a window size of 100kb. Insulation scores were then corrected across libraries using quantile normalization. Regions with differential insulation score were then identified using a linear regression model using the gender of the individual as a co-variate using the python package statsmodels (Seabold and Perktold 2010). Adjusted p-values were calculated using FDR correction (Benjamin-Hochberg) as implemented in the statsmodels package.

### Identification of loops and insulation bins altered in cell lines

To quantify how altered the chromatin conformation is in cell lines compared to primary cells we generated high quality Arima Hi-C libraries for two cell lines typically used as proxy for T cells. CD4^+^ T cells we used Jurkat E6-1 cells and for CD8^+^ T cells we used MyLa cells (Sigma-Aldrich, 95051033). Library generation was carried out using the same protocol as the primary cells. Sequencing was carried out to a final depth of 785 million reads for Jurkat cells and 385 million reads for MyLa cells and then processed using the same methodology as primary cells.

Loops and normalized counts were extracted as described in the previous sections. Because we had multiple replicates for the primary samples but only one replicate for the cell lines we had to test for differences in a different way. For each loop we calculated the mean and standard deviation of this loop from our dataset, and then tested if each sample was considered an outlier compared to the distribution of the counts for all samples. In practice this is calculated by calculating the z-score for each sample, then converting this to a p-value. The loop was considered an outlier if the unadjusted p-value was lower than 0.05.

Similarly, we carried out the same analysis for the insulation score. Insulation scores for all samples were pre-processed as described in the previous sections. Then for each genomic bin we calculated the mean and standard deviation of this bin in our dataset and then tested if each sample was considered an outlier in the same way as the loops.

### Loops correlation with gene expression and chromatin accessibility

Normalized counts for each loop, gene and ATAC-seq peak was extracted as previously described. Pearson’s correlation coefficient was calculated using the pearsonr function from scipy (Virtanen et al. 2020) between peak height or gene expression and interaction strength for each pairwise comparison for each sample. Because only loops that vary between the samples can show correlation, we only show the results for loops that are differentially interacting between CD4^+^ and CD8^+^ T cells (FDR<0.01). To limit computation time, we also only show results for chromosome 1, but similar results can be shown for any other chromosome. Additionally, we only test genes that are expressed in our samples (mean count > 5), loops that have a mean interaction strength of at least 10, and peaks with at least mean accessibility of 10 reads. The background used for peaks and genes all elements within 4 mb from the ends of the loop tested.

### Insulation score correlation with gene expression

Normalized insulation scores and counts for each gene were extracted as previously described. We then selected the genes that were differentially expressed (LFC>1, FDR<0.1) between CD4^+^ and CD8^+^ T cells and correlated their expression level with the insulation score of the bin which overlapped the promoter of the gene. Finally, the Pearson’s correlation coefficient was calculated using the pearsonr function from scipy (Virtanen et al. 2020).

### CTCF tracks

Imputed CTCF tracks for primary CD8^+^ and CD4^+^ T cells were obtained from ENCODE (ENCFF148DDI and ENCFF811QTU). These have been imputed using a multi-scale deep tensor factorization method, Avocado (Schreiber et al. 2020a, 2020b). To obtain peaks, we applied a threshold to the signal of 5, corresponding to genome-wide significance of a p-value of 10^−5^. Overlaps with loops and SNPs were obtained using pybedtools (Dale et al. 2011) and bedtools v2.30.0 (Quinlan and Hall 2010).

### Identification of loops with allelic imbalance

To identify loops displaying allelic imbalance we first had to generate phased genotypes for each individual. To obtain genotypes we first mapped all Hi-C reads for each individual to the GRCh38 genome using bwa-mem v0.7.17 using the settings -SP5M. Genotypes were then called using GLIMPSE v1.1.1 (Rubinacci et al. 2021) and the 1000 genomes phase 3 reference, making sure that all known restriction sites were excluded from the first step of the genotype calling. Genotype phasing was then carried out using the integrated phasing pipeline (Bansal 2019) which integrates population phasing using SHAPEIT2 (Delaneau et al. 2014) with reads phasing using the Hi-C using HAPCUT2 (Edge et al. 2017), resulting in around 2 million heterozygous SNPs phased per sample. We then reprocessed all Hi-C reads using Hi-C pro to a masked GRCh38 genome (all known SNPs masked with Ns) to reduce mapping bias. Aligned reads for each library were then split using SNPsplit v0.5.0 (Krueger and Andrews 2016) generating allele specific alignments. Counts for each allele for each significant loop (with a slop of 10kb) were then calculated using bedtools pair_to_pair (Quinlan and Hall 2010) and data was integrated in Python 3.9. Allelic imbalance of the reads for each loop that overlapped a SNP was tested using a one-sided binomial test (each SNP was tested twice for both directions separately) as implemented in scipy (Virtanen et al. 2020) for each sample which was heterozygous for that SNP. In addition, we filtered SNP/loop pairs so that at least 2 samples were heterozygous and phased for the SNP, at least 10 reads were present for each SNP and at least 1 read for each allele had to be present. All the resulting p-values were then merged using the Firsher’s method for meta-analysis as implemented in scipy (separately for each directionality). Resulting p-values were then corrected using Benjamini-Hochberg method as implemented in statsmodels (Seabold and Perktold 2010).

### Identification of variants with allelic imbalance in ATAC-seq

To identify variants displaying allelic imbalance in chromatin accessibility we first reprocessed all samples by mapping them to a masked GRCh38 genome (all known SNPs masked with Ns) to reduce mapping bias. For each sample we then identified the sites which contain variants using the bcftools v1.15.1 commands mpileup and call. Specifically, we used the following settings: mpileup -q 10 -I -E -a ‘FORMAT/DP,FORMAT/AD’ --ignore-RG - d 10000, call -Aim -C alleles. Additionally, we supplied the GRCh38 fasta references as well as the 1000 genomes sites, to limit the calling to known variants. Next, we filtered variants that had a read depth of at least 10 and had at least 2 reads from each allele or 5% of the reads, whichever was higher (REF and ALT). Read depth for each allele was extracted from the INFO field which contains the number of high-quality reads for the REF and the ATL alleles. Allelic imbalance for each SNP was tested using a one-sided binomial test (each SNP was tested twice for both directions separately) as implemented in scipy (Virtanen et al. 2020) for each sample. All the resulting p-values were then merged using the Firsher’s method for meta-analysis as implemented in scipy (separately for each directionality). Resulting p-values were then corrected using Benjamini-Hochberg method as implemented in statsmodels (Seabold and Perktold 2010).

Overlap with caQTL was done by first selecting the caQTLs for which the variant is directly overlapping the target ATAC-peak. Then overlaps were carried out by matching the rsID of the variant. Finally, we compared the directionality of the allelic imbalance with the slope of the caQTL for the variants that were significant both for allelic imbalance and for caQTL. Of note, a partial explanation for the limited overlap between caQTLs and allelic imbalance SNPs is the fundamental difference in the processing of the data. In particular, we found the following limitations:

- To compare read counts for ATAC-peaks across samples, a consensus peak set needs to be generated first. This limits the extent of the size of a promoter or enhancer to 500bp. Moreover, multiple enhancers within a certain size will be merged into one enhancer. We find that in some cases, we can split larger enhancers into multiple smaller sections, and these would correlate with genotype, whilst the overall called peak would not.
- Allelic imbalance can only be calculated if there are enough reads directly overlapping a SNP to support a heterozygous call.
- However, allelic imbalance intrinsically corrects for batch effects and other biological variability, as the imbalance is called from within the same sample.

### Genotyping array

Genotyping was carried out using the Illumina Infinium HumanCoreExome 24 BeadChip kit (Illumina, San Diego, California, USA). 250 ng of DNA was used, according to the manufacturer’s guidance. Genotype calling was carried out using GenomeStudio software (Illumina, San Diego, California, USA). Standard QC was conducted on each individual array using PLINK v1.9 (Purcell et al. 2007): SNPs and samples were excluded if there was >2% missing data, and SNPs with MAF□<□0.01 and Hardy Weinberg Equilibrium (HWE) p□<□1□×□10−4 were also excluded. Population stratification adjustment was done using HapMap 3 reference panel to determine genetic ancestry of each individual, followed by Principal Component Analysis (PCA) analysis. Full genotype imputation was carried out using HRC v1.1 reference panel on the Michigan Imputation Server (Das et al. 2016). Following imputation SNPs with imputation r2 score lower than 0.5 were removed.

### QTL analysis

Quantitative Trait Locus (QTL) analysis was performed to investigate the genetic basis of molecular phenotypes. The cis QTL analysis was carried out using QTLtools v1.3.1 (Delaneau et al. 2017), a versatile software that allows the use of arbitrary molecular phenotypes. We utilized genotypes called from the Hi-C data and the ATAC-seq data due to their lower error rates and fewer missing samples compared to those imputed from the genotyping array, although results were similar to the ones done using the array genotypes. Variants were filtered based on a minor allele frequency (MAF) greater than 0.05 in our samples.

For each molecular phenotype, samples that were identified as outliers through principal component analysis (PCA) were eliminated for each technique and condition. Four types of molecular phenotypes were investigated: gene expression QTL, chromatin accessibility QTL, insulation score QTL, and loop QTL.

Gene expression QTL analysis was conducted using gene expression data normalized with DESeq2 (Love et al. 2014) and log2 transformed. Chromatin accessibility QTL analysis was performed using counts from DiffBind (Stark and Brown 2011), which were subsequently quantile normalized and log2 transformed. Insulation score QTL analysis was conducted using insulation scores calculated as described previously and quantile normalized. Finally, loop QTL analysis was performed using loop counts extracted as described previously, followed by log2 transformation.

All QTL analyses were executed using the --normal parameter and tested regions within 1mb of the variant. The number of PCA components to be used as covariates for genotype and phenotype for each modality was determined by conducting a permutation pass on chromosome 1. The combination that yielded the most significant QTLs was then selected for further analysis. To calculate corrected p-values, we performed a permutation pass with 1000 permutations. However, corrected p-values are only calculated on the top hit. We find that for all our modalities an equivalent uncorrected p-value to a corrected p-value of 0.1 is approximately 1*10-4. For the calculation of the overlaps between the modalities, we decided to use this latter p-value as the top hit would rarely overlap between the different modalities.

For the overlaps between the same modality, we merged based on selecting one condition (CD4 or CD8) as the reference, which is filtered by corrected p-value of 0.1. using this as the lead signal for that eQTL we then searched the other condition for the same variant-phenotype pair at an uncorrected p-value of 1*10-4. We find that the concordance of the directionality is maintained at an uncorrected p-value of 1*10-3. For overlaps between eQTLs and caQTLs, we matched the ATAC-seq peaks that overlapped the TSS sites for the genes in the eQTL dataset. For overlaps with insQTLs we matched the insulation score bin to the position of the TSS or the ATAC-peak. For overlaps with loopQTLs we matched the TSS and the ATAC-seq peaks that overlapped either within the loop anchors or within the actual loop anchors, based on the description in the main text. When reporting results, we ran both CD8^+^ and CD4^+^ separately, then reported the average result between the two.

Since our dataset is relatively small compared to other publicly available eQTL datasets, we have integrated the eQTLgen dataset (Võsa et al. 2021) when discussing our results, where stated in the text.

### GWAS datasets and linkage disequilibrium SNPs identification

Leads SNPs for selected GWAS studies were downloaded as follows: PsA (Soomro et al. 2022), Ps (Tsoi et al. 2017), SSc (López-Isac et al. 2019), atopic dermatitis (Paternoster et al. 2015) and rheumatoid arthritis (Ishigaki et al. 2022), JIA (López-Isac et al. 2021).

SNPs in high linkage disequilibrium (R^2 > 0.8) with the lead SNPs were identified using plink v1.90b3.39 on the 1000 genomes data v3 with population set to EUR.

### Generation of enhancer deletion mouse

Two sgRNA for the upstream cut site (g483 – ggttacagtccgttaccgcc, g484 – tacgttcttcctggcggtaa) and two sgRHA downstream (g324 – gtgtgacatgagttgtcact, g325 – catgagttgtcactgggatt) were used as guides for the CRISPR-Cas9 deletion system. sgRNA were designed with stringent criteria for off target predictions (guides with mismatch (MM) of 0, 1 or 2 for elsewhere in the genome were discounted, and MM3 were tolerated if predicted off targets were not exonic according to http://www.sanger.ac.uk/htgt/wge/). The sgRNA were purchased as full length Alt-R sgRNA (IDT; Coralville, USA) and resupsended in sterile, RNase free injection buffer (TrisHCl 1 mM, pH 7.5, EDTA 0.1 mM). 1 ug each sgRNA were combined with 1 ug EnGen Cas9 (New England Biolabs). A final embryo injection mix (concentrations; each sgRNA 20 ng/ul, Cas9 protein 80 ng/ul) was directly microinjected into C57BL6/J (Envigo) zygote pronuclei using standard protocols. Zygotes were cultured overnight and the resulting 2 cell embryos surgically implanted into the oviduct of day 0.5 post-coitum pseudopregnant mice. Potential founder mice were identified by extraction of genomic DNA from ear clips, followed by PCR using primers that, upon an NHEJ mediated deletion event, are brought into proximity and generate an amplicon (Geno F1 GCTCTGCCACACTTGCTTAG, Geno R2 AACTGCACACGTGAACATGT). Sanger sequencing of the amplicon and alignment to the mouse genome verified the enhancer deletion in each pup.

## DATA AVAILABILITY STATEMENT

All processed data, differential gene expression and chromatin accessibility analysis, QTLs and allelic imbalance datasets are available online at:

http://bartzabel.ls.manchester.ac.uk/orozcolab/SNP2Mechanism/

All the code to reproduce the results presented here is available in the same webpage and on the GitHub repository:

https://github.com/ChenfuShi/PsA_cleaned_analysis

Precomputed Hi-C correlation maps with gene expression, chromatin accessibility and various variants are available at:

http://bartzabel.ls.manchester.ac.uk/orozcolab/SNP2Mechanism/PsA_output_hic_plots/main.html

Raw reads are available upon reasonable request.

## ACKNOWLEDGEMENTS

The authors would like to acknowledge the assistance given by IT Services and the use of the Computational Shared Facility at The University of Manchester, the study coordinators and the research nurses, the flow cytometry facility and the patients and healthy volunteers who contributed their time and donated the samples.

This work was funded by the Wellcome Trust (award references 207491/Z/17/Z and 215207/Z/19/Z), Versus Arthritis (award reference 21754), NIHR Manchester Biomedical Research Centre and the Medical Research Council (award reference MR/N00017X/1).

## CONTRIBUTIONS

Example: DZ, SR, AF, JD, CF, CW, RH, ER and MG collected and processed the samples. CS, DZ, SR, AF, JD, ER, MG and AA designed and performed the experiments. CS and CY analysed the data. DP, PM, SE, JB, AB, PH, MR and GO acquired the funding and oversaw the project. CS and GO conceived the work, generated the figures, and wrote the manuscript. All authors have read and approved the final manuscript.

---

**Supplementary Figure 1.** A) UMAP of RNA-seq data, counts from DESeq2, symbol shape is sex of individual B) UMAP of the ATAC-seq data, counts from DiffBind, symbol shape is sex of individual. C) UMAP of the Hi-C loops data, symbol shape is sex of individual. D) MDS of HiCRep analysis of the Hi-C data, symbol shape is sex of individual.

**Supplementary Figure 2.** A) APA plot of the loops that overlap CTCF at both ends. Numbers at bottom left corner and upper right corner indicate the ratio between the central pixel and the mean of the pixels at those corners. All loops and Hi-C map are from merged CD8^+^ T cells Hi-C. B) APA plot of the loops that overlap CTCF at least at one end. C) APA plot of the loops that do not overlap CTCF. D) Boxplot with number of insulation score bins that were considered outliers, per sample. MyLa and Jurkat cells are highlighted. E) boxplot with number of loops that were considered outliers, per sample. MyLa and Jurkat cells are highlighted.

**Supplementary Figure 3.** A) Hi-C maps from the *ANKRD44* locus, showing differences in chromatin conformation between Jurkat CD4^+^ T cells and primary CD4^+^ T cells. B) Hi-C maps from the *KLRF1* locus, showing differences in chromatin conformation between Jurkat CD4^+^ T cells and primary CD4^+^ T cells.

**Supplementary Figure 4.** A) Hi-C maps from the *ITK* locus, showing differences in chromatin conformation between Jurkat CD4^+^ T cells and primary CD4^+^ T cells. B) Hi-C maps from the *GZMB* locus, showing differences in chromatin conformation between Jurkat CD4^+^ T cells and primary CD4^+^ T cells.

**Supplementary Figure 5.** A) Hi-C maps from the *SLC22A5* PsA GWAS locus, showing differences in chromatin conformation between MyLa CD8^+^ T cells and primary CD8^+^ T cells. B) Hi-C maps from the *FYN* PsA GWAS locus, showing differences in chromatin conformation between Jurkat CD4^+^ T cells and primary CD4^+^ T cells.

**Supplementary Figure 6.** A) Accessibility of CTCF site located at chr19:39440211 vs the genotype of rs2353678. B) Visualization of the region surrounding rs2353678. Top: ATAC-seq peaks. Intensity of the colour indicates the average peak-height of the peak, whilst position across the Y axis indicates the pearson’s correlation with the genotype of rs2353678. Middle: Genes from Gencode v29. The red dots indicate the transcription start sites. Bottom left: Correlation between the Hi-C contacts and the genotype of rs2353678. Bottom right: Merged Hi-C map.

**Supplementary Figure 7.** A) Region view from ensembl genome browser with homology between *Homo Sapiens* and *Mus Musculus* for the *BUB1, ACOXL, BCL2L11* region. B) same as before but zoomed in view of the regulatory region, with the 3 chromatin accessibility peaks that were deleted in mouse.

## REFERENCES

1. Alasoo K, Rodrigues J, Mukhopadhyay S, Knights AJ, Mann AL, Kundu K, et al. Shared genetic effects on chromatin and gene expression indicate a role for enhancer priming in immune response. Nat Genet [Internet]. 2018;50(3):424–31. Available from: http://dx.doi.org/10.1038/s41588-018-0046-7

2. Aljahani A, Hua P, Karpinska MA, Quililan K, Davies JOJ, Oudelaar AM. Analysis of sub-kilobase chromatin topology reveals nano-scale regulatory interactions with variable dependence on cohesin and CTCF. Nat Commun 2022 131 [Internet]. 2022 Apr 19 [cited 2022 Apr 21];13(1):1–13. Available from: https://www.nature.com/articles/s41467-022-29696-5

3. Amanda Ackermann. ATAC-seq protocol [Internet]. Kaestner Lab. 2019 [cited 2022 May 13]. Available from: https://www.med.upenn.edu/kaestnerlab/assets/user-content/documents/ATAC-seq-Protocol-(Omni)-Kaestner-Lab.pdf

4. An L, Yang T, Yang J, Nuebler J, Xiang G, Hardison RC, et al. OnTAD: Hierarchical domain structure reveals the divergence of activity among TADs and boundaries. Genome Biol [Internet]. 2019 Dec 18 [cited 2023 Jul 7];20(1):1–16. Available from: https://genomebiology.biomedcentral.com/articles/10.1186/s13059-019-1893-y

5. Ardakany AR, Gezer HT, Lonardi S, Ay F. Mustache: Multi-scale Detection of Chromatin Loops from Hi-C and Micro-C Maps using Scale-Space Representation. bioRxiv. 2020 Feb 26;2020.02.24.963579.

6. Argyriou A, Wadsworth MH, Lendvai A, Christensen SM, Hensvold AH, Gerstner C, et al. Single cell sequencing identifies clonally expanded synovial CD4+ TPH cells expressing GPR56 in rheumatoid arthritis. Nat Commun 2022 131 [Internet]. 2022 Jul 13 [cited 2023 Jul 19];13(1):1–13. Available from: https://www.nature.com/articles/s41467-022-31519-6

7. Bansal V. Integrating read-based and population-based phasing for dense and accurate haplotyping of individual genomes. Bioinformatics [Internet]. 2019 Jul 15 [cited 2022 Jul 27];35(14):i242–8. Available from: https://academic.oup.com/bioinformatics/article/35/14/i242/5529122

8. Bhattacharyya S, Chandra V, Vijayanand P, Ay F. Identification of significant chromatin contacts from HiChIP data by FitHiChIP. Nat Commun. 2019 Dec;10(1).

9. Boix CA, James BT, Park YP, Meuleman W, Kellis M. Regulatory genomic circuitry of human disease loci by integrative epigenomics. Nature [Internet]. 2021 Feb 3 [cited 2021 Feb 4];590:1–8. Available from: http://www.nature.com/articles/s41586-020-03145-z

10. Bouillet P, Metcalf D, Huang DCS, Tarlinton DM, Kay TWH, Kontgen F, et al. Proapoptotic Bcl-2 Relative Bim Required for Certain Apoptotic Responses, Leukocyte Homeostasis, and to Preclude Autoimmunity. Science (80-) [Internet]. 1999 Nov 26 [cited 2023 Jul 14];286(5445):1735–8. Available from: https://www.science.org/doi/10.1126/science.286.5445.1735

11. Buniello A, Macarthur JAL, Cerezo M, Harris LW, Hayhurst J, Malangone C, et al. The NHGRI-EBI GWAS Catalog of published genome-wide association studies, targeted arrays and summary statistics 2019. Nucleic Acids Res [Internet]. 2019 Jan 8 [cited 2020 Oct 1];47(D1):D1005–12. Available from: https://pubmed.ncbi.nlm.nih.gov/30445434/

12. Cairns J, Freire-Pritchett P, Wingett SW, Várnai C, Dimond A, Plagnol V, et al. CHiCAGO: robust detection of DNA looping interactions in Capture Hi-C data. Genome Biol [Internet]. 2016 Dec 15 [cited 2018 Apr 24];17(1):127. Available from: http://genomebiology.biomedcentral.com/articles/10.1186/s13059-016-0992-2

13. Cano-Gamez E, Trynka G. From GWAS to Function: Using Functional Genomics to Identify the Mechanisms Underlying Complex Diseases. Front Genet. 2020 May 13;11:505357.

14. Chen S, Zhou Y, Chen Y, Gu J. fastp: an ultra-fast all-in-one FASTQ preprocessor. Bioinformatics [Internet]. 2018 Sep 1 [cited 2019 Nov 19];34(17):i884–90. Available from: https://academic.oup.com/bioinformatics/article/34/17/i884/5093234

15. Choy M-K, Javierre BM, Williams SG, Baross SL, Liu Y, Wingett SW, et al. Promoter interactome of human embryonic stem cell-derived cardiomyocytes connects GWAS regions to cardiac gene networks. Nat Commun [Internet]. 2018 Dec 28 [cited 2019 Mar 6];9(1):2526. Available from: http://www.nature.com/articles/s41467-018-04931-0

16. Corces MR, Trevino AE, Hamilton EG, Greenside PG, Sinnott-Armstrong NA, Vesuna S, et al. An improved ATAC-seq protocol reduces background and enables interrogation of frozen tissues. Nat Methods 2017 1410 [Internet]. 2017 Aug 28 [cited 2022 May 12];14(10):959–62. Available from: https://www.nature.com/articles/nmeth.4396

17. Dale RK, Pedersen BS, Quinlan AR. Pybedtools: A flexible Python library for manipulating genomic datasets and annotations. Bioinformatics [Internet]. 2011 Dec 15 [cited 2020 Oct 1];27(24):3423–4. Available from: http://pypi.python.org/pypi/pybedtools.

18. Das S, Forer L, Schönherr S, Sidore C, Locke AE, Kwong A, et al. Next-generation genotype imputation service and methods. Nat Genet [Internet]. 2016 Oct 1 [cited 2023 Jul 6];48(10):1284–7. Available from: https://pubmed.ncbi.nlm.nih.gov/27571263/

19. Delaneau O, Marchini J, McVeanh GA, Donnelly P, Lunter G, Marchini JL, et al. Integrating sequence and array data to create an improved 1000 Genomes Project haplotype reference panel. Nat Commun 2014 51 [Internet]. 2014 Jun 13 [cited 2022 Jul 27];5(1):1–9. Available from: https://www.nature.com/articles/ncomms4934

20. Delaneau O, Ongen H, Brown AA, Fort A, Panousis NI, Dermitzakis ET. A complete tool set for molecular QTL discovery and analysis. Nat Commun 2017 81 [Internet]. 2017 May 18 [cited 2023 May 5];8(1):1–7. Available from: https://www.nature.com/articles/ncomms15452

21. Delaneau O, Zazhytska M, Borel C, Giannuzzi G, Rey G, Howald C, et al. Chromatin three-dimensional interactions mediate genetic effects on gene expression. Science (80-). 2019 May 3;364(6439).

22. Dixon JR, Jung I, Selvaraj S, Shen Y, Antosiewicz-Bourget JE, Lee AY, et al. Chromatin architecture reorganization during stem cell differentiation. Nature [Internet]. 2015 Feb 19 [cited 2018 Sep 20];518(7539):331–6. Available from: http://www.nature.com/articles/nature14222

23. Edge P, Bafna V, Bansal V. HapCUT2: robust and accurate haplotype assembly for diverse sequencing technologies. Genome Res [Internet]. 2017 Dec 9 [cited 2019 Sep 9];27(5):801–12. Available from: http://www.ncbi.nlm.nih.gov/pubmed/27940952

24. Farh KK-H, Marson A, Zhu J, Kleinewietfeld M, Housley WJ, Beik S, et al. Genetic and epigenetic fine mapping of causal autoimmune disease variants. Nature [Internet]. 2015 Feb [cited 2018 Jun 12];518(7539):337–43. Available from: http://www.ncbi.nlm.nih.gov/pubmed/25363779

25. Ferreira MAR, Mathur R, Vonk JM, Szwajda A, Brumpton B, Granell R, et al. Genetic Architectures of Childhood- and Adult-Onset Asthma Are Partly Distinct. Am J Hum Genet [Internet]. 2019 Apr 4 [cited 2023 Jul 19];104(4):665–84. Available from: https://pubmed.ncbi.nlm.nih.gov/30929738/

26. Finucane HK, Bulik-Sullivan B, Gusev A, Trynka G, Reshef Y, Loh PR, et al. Partitioning heritability by functional annotation using genome-wide association summary statistics. Nat Genet 2015 4711 [Internet]. 2015 Sep 28 [cited 2022 Mar 25];47(11):1228–35. Available from: https://www.nature.com/articles/ng.3404

27. Fort A, Panousis NI, Garieri M, Antonarakis SE, Lappalainen T, Dermitzakis ET, et al. MBV: a method to solve sample mislabeling and detect technical bias in large combined genotype and sequencing assay datasets. Bioinformatics [Internet]. 2017 Jun 15 [cited 2023 May 5];33(12):1895–7. Available from: https://academic.oup.com/bioinformatics/article/33/12/1895/2982050

28. Fullwood MJ, Han Y, Wei CL, Ruan X, Ruan Y. Chromatin interaction analysis using paired-end tag sequencing. Vol. CHAPTER 21, Current Protocols in Molecular Biology. NIH Public Access; 2010. p. Unit.

29. Ge X, Frank-Bertoncelj M, Klein K, McGovern A, Kuret T, Houtman M, et al. Functional genomics atlas of synovial fibroblasts defining rheumatoid arthritis heritability. Genome Biol [Internet]. 2021 Dec 1 [cited 2022 Feb 24];22(1):1–39. Available from: https://genomebiology.biomedcentral.com/articles/10.1186/s13059-021-02460-6

30. Goel VY, Huseyin MK, Hansen AS. Region Capture Micro-C reveals coalescence of enhancers and promoters into nested microcompartments. Nat Genet [Internet]. 2023 Jun 8 [cited 2023 Jul 4];55(6):1048–56. Available from: https://pubmed.ncbi.nlm.nih.gov/37157000/

31. González-Serna D, Shi C, Kerick M, Hankinson J, Ding J, McGovern A, et al. Identification of Mechanisms by Which Genetic Susceptibility Loci Influence Systemic Sclerosis Risk Using Functional Genomics in Primary T Cells and Monocytes. Arthritis Rheumatol (Hoboken, NJ) [Internet]. 2023 Jun 1 [cited 2023 Jul 11];75(6):1007–20. Available from: https://pubmed.ncbi.nlm.nih.gov/36281738/

32. Gorkin DU, Qiu Y, Hu M, Fletez-Brant K, Liu T, Schmitt AD, et al. Common DNA sequence variation influences 3-dimensional conformation of the human genome. Genome Biol [Internet]. 2019 Dec 28 [cited 2019 Nov 29];20(1):255. Available from: https://genomebiology.biomedcentral.com/articles/10.1186/s13059-019-1855-4

33. Greenwald WW, Li H, Benaglio P, Jakubosky D, Matsui H, Schmitt A, et al. Subtle changes in chromatin loop contact propensity are associated with differential gene regulation and expression. Nat Commun [Internet]. 2019 Dec 5 [cited 2019 Mar 15];10(1):1054. Available from: http://www.nature.com/articles/s41467-019-08940-5

34. Hastings N, Agaba M, Tocher DR, Leaver MJ, Dick JR, Sargent JR, et al. A vertebrate fatty acid desaturase with Δ5 and Δ6 activities. Proc Natl Acad Sci U S A [Internet]. 2001 Dec 4 [cited 2023 Jul 19];98(25):14304–9. Available from: https://www.pnas.org/doi/abs/10.1073/pnas.251516598

35. Hojo MA, Masuda K, Hojo H, Nagahata Y, Yasuda K, Ohara D, et al. Identification of a genomic enhancer that enforces proper apoptosis induction in thymic negative selection. Nat Commun 2019 101 [Internet]. 2019 Jun 13 [cited 2023 Jun 27];10(1):1–15. Available from: https://www.nature.com/articles/s41467-019-10525-1

36. Hsieh THS, Cattoglio C, Slobodyanyuk E, Hansen AS, Rando OJ, Tjian R, et al. Resolving the 3D Landscape of Transcription-Linked Mammalian Chromatin Folding. Mol Cell. 2020 May 7;78(3):539–553.e8.

37. Hua P, Badat M, Hanssen LLP, Hentges LD, Crump N, Downes DJ, et al. Defining genome architecture at base-pair resolution. Nat 2021 5957865 [Internet]. 2021 Jun 9 [cited 2022 Feb 8];595(7865):125–9. Available from: https://www.nature.com/articles/s41586-021-03639-4

38. Institute B. Picard toolkit. Broad Institute, GitHub Repos [Internet]. 2019 [cited 2022 May 12]; Available from: https://broadinstitute.github.io/picard/

39. Iqbal MM, Serralha M, Kaur P, Martino D. Mapping the landscape of chromatin dynamics during naïve CD4+ T-cell activation. Sci Rep. 2021 Dec 1;11(1).

40. Ishigaki K, Sakaue S, Terao C, Luo Y, Sonehara K, Yamaguchi K, et al. Multi-ancestry genome-wide association analyses identify novel genetic mechanisms in rheumatoid arthritis. Nat Genet 2022 5411 [Internet]. 2022 Nov 4 [cited 2023 Apr 21];54(11):1640–51. Available from: https://www.nature.com/articles/s41588-022-01213-w

41. Javierre BM, Burren OS, Wilder SP, Kreuzhuber R, Hill SM, Sewitz S, et al. Lineage-Specific Genome Architecture Links Enhancers and Non-coding Disease Variants to Target Gene Promoters. Cell [Internet]. 2016 Nov 17 [cited 2018 Jun 26];167(5):1369–1384.e19. Available from: http://www.ncbi.nlm.nih.gov/pubmed/27863249

42. King EA, Wade Davis J, Degner JF. Are drug targets with genetic support twice as likely to be approved? Revised estimates of the impact of genetic support for drug mechanisms on the probability of drug approval. PLOS Genet [Internet]. 2019 [cited 2022 Apr 7];15(12):e1008489. Available from: https://journals.plos.org/plosgenetics/article?id=10.1371/journal.pgen.1008489

43. Kostoglou-Athanassiou I, Athanassiou L, Athanassiou P. The Effect of Omega-3 Fatty Acids on Rheumatoid Arthritis. Mediterr J Rheumatol [Internet]. 2020 [cited 2023 Jul 19];31(2):190. Available from: /pmc/articles/PMC7362115/

44. Krietenstein N, Abraham S, Venev S V., Abdennur N, Gibcus J, Hsieh THS, et al. Ultrastructural Details of Mammalian Chromosome Architecture. Mol Cell. 2020;78(3):554–565.e7.

45. Krueger F, Andrews SR. SNPsplit: Allele-specific splitting of alignments between genomes with known SNP genotypes. F1000Research [Internet]. 2016 [cited 2022 Jul 27];5. Available from: https://pubmed.ncbi.nlm.nih.gov/27429743/

46. Kundaje A, Meuleman W, Ernst J, Bilenky M, Yen A, Heravi-Moussavi A, et al. Integrative analysis of 111 reference human epigenomes. Nature [Internet]. 2015 Feb 19 [cited 2018 Jul 7];518(7539):317–30. Available from: http://www.nature.com/articles/nature14248

47. Langmead B, Salzberg SL. Fast gapped-read alignment with Bowtie 2. Nat Methods [Internet]. 2012 Apr 4 [cited 2018 Jul 7];9(4):357–9. Available from: http://www.nature.com/articles/nmeth.1923

48. Lin D, Sanders J, Noble WS, Allen PG. HiCRep.py: fast comparison of Hi-C contact matrices in Python. Bioinformatics [Internet]. 2021 Sep 29 [cited 2022 Mar 17];37(18):2996–7. Available from: https://academic.oup.com/bioinformatics/article/37/18/2996/6133255

49. López-Isac E, Acosta-Herrera M, Kerick M, Assassi S, Satpathy AT, Granja J, et al. GWAS for systemic sclerosis identifies multiple risk loci and highlights fibrotic and vasculopathy pathways. Nat Commun. 2019 Dec 1;10(1).

50. López-Isac E, Smith SL, Marion MC, Wood A, Sudman M, Yarwood A, et al. Combined genetic analysis of juvenile idiopathic arthritis clinical subtypes identifies novel risk loci, target genes and key regulatory mechanisms. Ann Rheum Dis [Internet]. 2021 Mar 1 [cited 2022 Feb 24];80(3):321–8. Available from: https://ard.bmj.com/content/80/3/321

51. Love MI, Huber W, Anders S. Moderated estimation of fold change and dispersion for RNA-seq data with DESeq2. Genome Biol [Internet]. 2014 Dec 5 [cited 2018 Jul 15];15(12):550. Available from: http://genomebiology.biomedcentral.com/articles/10.1186/s13059-014-0550-8

52. Martin P, McGovern A, Orozco G, Duffus K, Yarwood A, Schoenfelder S, et al. Capture Hi-C reveals novel candidate genes and complex long-range interactions with related autoimmune risk loci. Nat Commun [Internet]. 2015;6:1–7. Available from: http://dx.doi.org/10.1038/ncomms10069

53. McGovern A, Schoenfelder S, Martin P, Massey J, Duffus K, Plant D, et al. Capture Hi-C identifies a novel causal gene, IL20RA, in the pan-autoimmune genetic susceptibility region 6q23. Genome Biol [Internet]. 2016;17(1). Available from: http://dx.doi.org/10.1186/s13059-016-1078-x

54. McInnes L, Healy J, Melville J. UMAP: Uniform Manifold Approximation and Projection for Dimension Reduction. 2018 Feb 9 [cited 2022 May 12]; Available from: https://arxiv.org/abs/1802.03426v3

55. Mifsud B, Tavares-Cadete F, Young AN, Sugar R, Schoenfelder S, Ferreira L, et al. Mapping long-range promoter contacts in human cells with high-resolution capture Hi-C. Nat Genet [Internet]. 2015 Jun 4 [cited 2018 Oct 3];47(6):598–606. Available from: http://www.nature.com/articles/ng.3286

56. Miguel-Escalada I, Bonàs-Guarch S, Cebola I, Ponsa-Cobas J, Mendieta-Esteban J, Atla G, et al. Human pancreatic islet three-dimensional chromatin architecture provides insights into the genetics of type 2 diabetes. Nat Genet [Internet]. 2019 Jul 28 [cited 2019 Jul 10];51(7):1137–48. Available from: http://www.nature.com/articles/s41588-019-0457-0

57. Montefiori LE, Sobreira DR, Sakabe NJ, Aneas I, Joslin AC, Hansen GT, et al. A promoter interaction map for cardiovascular disease genetics. Elife [Internet]. 2018 Jul 10 [cited 2018 Nov 28];7. Available from: http://www.ncbi.nlm.nih.gov/pubmed/29988018

58. Mumbach MR, Satpathy AT, Boyle EA, Dai C, Gowen BG, Cho SW, et al. Enhancer connectome in primary human cells identifies target genes of disease-associated DNA elements. Nat Genet [Internet]. 2017 Sep 25 [cited 2018 Jun 15];49(11):1602–12. Available from: http://www.nature.com/doifinder/10.1038/ng.3963

59. Nolis IK, McKay DJ, Mantouvalou E, Lomvardas S, Merika M, Thanos D. Transcription factors mediate long-range enhancer-promoter interactions. Proc Natl Acad Sci U S A [Internet]. 2009 Dec 1 [cited 2018 Nov 26];106(48):20222–7. Available from: http://www.ncbi.nlm.nih.gov/pubmed/19923429

60. O’Rielly DD, Rahman P. Genetics of susceptibility and treatment response in psoriatic arthritis. Nat Rev Rheumatol [Internet]. 2011 Dec 8 [cited 2018 Sep 1];7(12):718–32. Available from: http://www.nature.com/articles/nrrheum.2011.169

61. Orozco G. Fine mapping with epigenetic information and 3D structure. Semin Immunopathol [Internet]. 2022 Jan 1 [cited 2023 Jul 7];44(1):115–25. Available from: https://pubmed.ncbi.nlm.nih.gov/35022890/

62. Orozco G, Schoenfelder S, Walker N, Eyre S, Fraser P. 3D genome organization links non-coding disease-associated variants to genes. Front Cell Dev Biol. 2022 Oct 20;10:995388.

63. Paternoster L, Standl M, Waage J, Baurecht H, Hotze M, Strachan DP, et al. Multi-ancestry genome-wide association study of 21,000 cases and 95,000 controls identifies new risk loci for atopic dermatitis. Nat Genet. 2015 Dec 1;47(12):1449–56.

64. Patro R, Duggal G, Love MI, Irizarry RA, Kingsford C. Salmon provides fast and bias-aware quantification of transcript expression. Nat Methods [Internet]. 2017 Apr [cited 2018 Jul 15];14(4):417–9. Available from: http://www.ncbi.nlm.nih.gov/pubmed/28263959

65. Purcell S, Neale B, Todd-Brown K, Thomas L, Ferreira MAR, Bender D, et al. PLINK: A Tool Set for Whole-Genome Association and Population-Based Linkage Analyses. Am J Hum Genet [Internet]. 2007 [cited 2023 Jul 6];81(3):559. Available from: /pmc/articles/PMC1950838/

66. Quinlan AR, Hall IM. BEDTools: a flexible suite of utilities for comparing genomic features. Bioinformatics [Internet]. 2010 Mar 15 [cited 2018 Jul 7];26(6):841–2. Available from: https://academic.oup.com/bioinformatics/article-lookup/doi/10.1093/bioinformatics/btq033

67. Ramírez F, Ryan DP, Grüning B, Bhardwaj V, Kilpert F, Richter AS, et al. deepTools2: a next generation web server for deep-sequencing data analysis. Nucleic Acids Res [Internet]. 2016 Jul 8 [cited 2018 Jul 7];44(W1):W160–5. Available from: https://academic.oup.com/nar/article-lookup/doi/10.1093/nar/gkw257

68. Rao SSP, Huntley MH, Durand NC, Stamenova EK, Bochkov ID, Robinson JT, et al. A 3D map of the human genome at kilobase resolution reveals principles of chromatin looping. Cell [Internet]. 2014 Dec 18 [cited 2018 Oct 1];159(7):1665–80. Available from: http://www.ncbi.nlm.nih.gov/pubmed/25497547

69. Ray-Jones H, Duffus K, McGovern A, Martin P, Shi C, Hankinson J, et al. Mapping DNA interaction landscapes in psoriasis susceptibility loci highlights KLF4 as a target gene in 9q31. BMC Biol [Internet]. 2020 May 4 [cited 2020 May 7];18(1):1–20. Available from: https://bmcbiol.biomedcentral.com/articles/10.1186/s12915-020-00779-3

70. Robertson CC, Inshaw JRJ, Onengut-Gumuscu S, Chen WM, Santa Cruz DF, Yang H, et al. Fine-mapping, trans-ancestral and genomic analyses identify causal variants, cells, genes and drug targets for type 1 diabetes. Nat Genet 2021 537 [Internet]. 2021 Jun 14 [cited 2023 Apr 21];53(7):962–71. Available from: https://www.nature.com/articles/s41588-021-00880-5

71. Rowley MJ, Corces VG. Organizational principles of 3D genome architecture. Nat Rev Genet [Internet]. 2018 Oct 26 [cited 2018 Oct 30];1. Available from: http://www.nature.com/articles/s41576-018-0060-8

72. Rubinacci S, Ribeiro DM, Hofmeister RJ, Delaneau O. Efficient phasing and imputation of low-coverage sequencing data using large reference panels. Nat Genet 2021 531 [Internet]. 2021 Jan 7 [cited 2022 Jul 27];53(1):120–6. Available from: https://www.nature.com/articles/s41588-020-00756-0

73. Sadowski M, Kraft A, Szalaj P, Wlasnowolski M, Tang Z, Ruan Y, et al. Spatial chromatin architecture alteration by structural variations in human genomes at the population scale. Genome Biol [Internet]. 2019 Jul 30 [cited 2023 May 10];20(1):1–27. Available from: https://genomebiology.biomedcentral.com/articles/10.1186/s13059-019-1728-x

74. Schmiedel BJ, Seumois G, Samaniego-Castruita D, Cayford J, Schulten V, Chavez L, et al. 17q21 asthma-risk variants switch CTCF binding and regulate IL-2 production by T cells. Nat Commun [Internet]. 2016 Nov 16 [cited 2023 Jul 4];7. Available from: /pmc/articles/PMC5116091/

75. Schmiedel BJ, Singh D, Madrigal A, Valdovino-Gonzalez AG, White BM, Zapardiel-Gonzalo J, et al. Impact of Genetic Polymorphisms on Human Immune Cell Gene Expression. Cell [Internet]. 2018 Nov 29 [cited 2018 Dec 3];175(6):1701–1715.e16. Available from: https://www.sciencedirect.com/science/article/pii/S009286741831331X

76. Schmitt AD, Hu M, Jung I, Lin Y, Barr CL, Ren B. A Compendium of Chromatin Contact Maps Reveals Spatially Active Regions in the Human Genome. CellReports [Internet]. 2016 [cited 2018 Sep 20];17:2042–59. Available from: http://www.cell.com/consortium/IHEC.http://dx.doi.org/10.1016/j.celrep.2016.10.061

77. Schreiber J, Bilmes J, Noble WS. Completing the ENCODE3 compendium yields accurate imputations across a variety of assays and human biosamples. Genome Biol [Internet]. 2020a Mar 30 [cited 2022 Mar 16];21(1):1–13. Available from: https://genomebiology.biomedcentral.com/articles/10.1186/s13059-020-01978-5

78. Schreiber J, Durham T, Bilmes J, Noble WS. Avocado: A multi-scale deep tensor factorization method learns a latent representation of the human epigenome. Genome Biol [Internet]. 2020b Mar 30 [cited 2022 May 12];21(1):1–18. Available from: https://genomebiology.biomedcentral.com/articles/10.1186/s13059-020-01977-6

79. Seabold S, Perktold J. statsmodels: Econometric and statistical modeling with python. In: 9th Python in Science Conference. 2010.

80. Servant N, Varoquaux N, Lajoie BR, Viara E, Chen C-J, Vert J-P, et al. HiC-Pro: an optimized and flexible pipeline for Hi-C data processing. Genome Biol [Internet]. 2015 Dec 1 [cited 2018 Oct 1];16(1):259. Available from: http://genomebiology.com/2015/16/1/259

81. Shi C, Ray-Jones H, Ding J, Duffus K, Fu Y, Gaddi VP, et al. Chromatin Looping Links Target Genes with Genetic Risk Loci for Dermatological Traits. J Invest Dermatol. 2021 Aug 1;141(8):1975–84.

82. Shlyueva D, Stampfel G, Stark A. Transcriptional enhancers: from properties to genome-wide predictions. Nat Rev Genet [Internet]. 2014 Apr 11 [cited 2018 Jun 19];15(4):272–86. Available from: http://www.nature.com/articles/nrg3682

83. Simeonov DR, Gowen BG, Boontanrart M, Roth TL, Gagnon JD, Mumbach MR, et al. Discovery of stimulation-responsive immune enhancers with CRISPR activation. Nature [Internet]. 2017;549(7670):111–5. Available from: http://dx.doi.org/10.1038/nature23875

84. Soomro M, Stadler M, Dand N, Bluett J, Jadon D, Jalali-najafabadi F, et al. Comparative genetic analysis of psoriatic arthritis and psoriasis for the discovery of genetic risk factors and risk prediction modelling. Arthritis Rheumatol [Internet]. 2022 May 4 [cited 2022 Jun 3]; Available from: https://onlinelibrary.wiley.com/doi/full/10.1002/art.42154

85. van Staalduinen J, van Staveren T, Grosveld F, Wendt KS. Live-cell imaging of chromatin contacts opens a new window into chromatin dynamics. Epigenetics Chromatin [Internet]. 2023 Dec 1 [cited 2023 Jul 12];16(1). Available from: /pmc/articles/PMC10288748/

86. Stark R, Brown G. DiffBind: Differential binding analysis of ChIP-Seq peak data [Internet]. 2011. Available from: http://bioconductor.org/packages/release/bioc/vignettes/DiffBind/inst/doc/DiffBind.pdf

87. Tsoi LC, Stuart PE, Tian C, Gudjonsson JE, Das S, Zawistowski M, et al. Large scale meta-analysis characterizes genetic architecture for common psoriasis associated variants. Nat Commun [Internet]. 2017 Aug 24 [cited 2019 Jun 19];8(1):15382. Available from: http://www.nature.com/articles/ncomms15382

88. Veale DJ, Fearon U. The pathogenesis of psoriatic arthritis. Lancet (London, England) [Internet]. 2018 Jun 2 [cited 2018 Sep 2];391(10136):2273–84. Available from: http://www.ncbi.nlm.nih.gov/pubmed/29893226

89. Venev S, Abdennur N, Goloborodko A, Flyamer I, Fudenberg G, Nuebler J, et al. open2c/cooltools: v0.5.1. 2022 Mar 2 [cited 2022 May 12]; Available from: https://zenodo.org/record/6324229

90. Verlaan DJ, Berlivet S, Hunninghake GM, Madore AM, Larivière M, Moussette S, et al. Allele-Specific Chromatin Remodeling in the ZPBP2/GSDMB/ORMDL3 Locus Associated with the Risk of Asthma and Autoimmune Disease. Am J Hum Genet. 2009 Sep 11;85(3):377–93.

91. Virtanen P, Gommers R, Oliphant TE, Haberland M, Reddy T, Cournapeau D, et al. SciPy 1.0: fundamental algorithms for scientific computing in Python. Nat Methods 2020 173 [Internet]. 2020 Feb 3 [cited 2023 Jul 12];17(3):261–72. Available from: https://www.nature.com/articles/s41592-019-0686-2

92. Võsa U, Claringbould A, Westra HJ, Bonder MJ, Deelen P, Zeng B, et al. Large-scale cis- and trans-eQTL analyses identify thousands of genetic loci and polygenic scores that regulate blood gene expression. Nat Genet. 2021;53(9):1300–10.

93. Xu W, Zhong Q, Lin D, Zuo Y, Dai J, Li G, et al. CoolBox: a flexible toolkit for visual analysis of genomics data. BMC Bioinformatics [Internet]. 2021 Dec 1 [cited 2022 May 13];22(1):1–9. Available from: https://bmcbioinformatics.biomedcentral.com/articles/10.1186/s12859-021-04408-w

94. Yang J, McGovern A, Martin P, Duffus K, Ge X, Zarrineh P, et al. Analysis of chromatin organization and gene expression in T cells identifies functional genes for rheumatoid arthritis. Nat Commun 2020 111 [Internet]. 2020 Sep 2 [cited 2022 Mar 25];11(1):1–13. Available from: https://www.nature.com/articles/s41467-020-18180-7

95. Yang T, Zhang F, Yardımcı GG, Song F, Hardison RC, Noble WS, et al. HiCRep: assessing the reproducibility of Hi-C data using a stratum-adjusted correlation coefficient. Genome Res [Internet]. 2017 Nov 1 [cited 2018 Sep 17];27(11):1939–49. Available from: http://www.ncbi.nlm.nih.gov/pubmed/28855260

96. Zhang Y, Liu T, Meyer CA, Eeckhoute J, Johnson DS, Bernstein BE, et al. Model-based Analysis of ChIP-Seq (MACS). Genome Biol [Internet]. 2008 Sep 17 [cited 2018 Jul 7];9(9):R137. Available from: http://genomebiology.biomedcentral.com/articles/10.1186/gb-2008-9-9-r137

