## Supplementary figures and images for "Multi-omics analysis in primary T cells elucidates mechanisms behind disease associated genetic loci"

### Figure s1

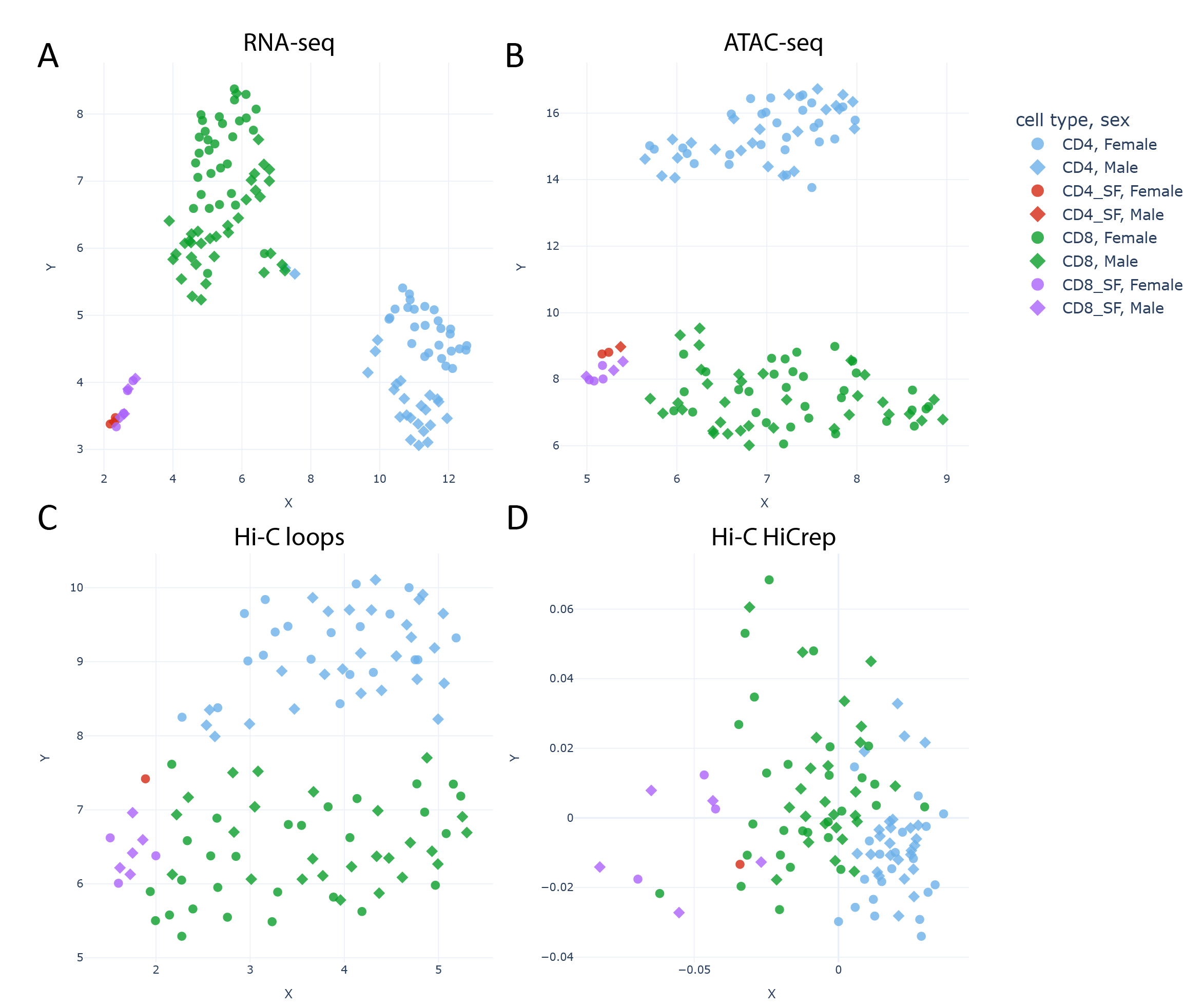

### Figure s2

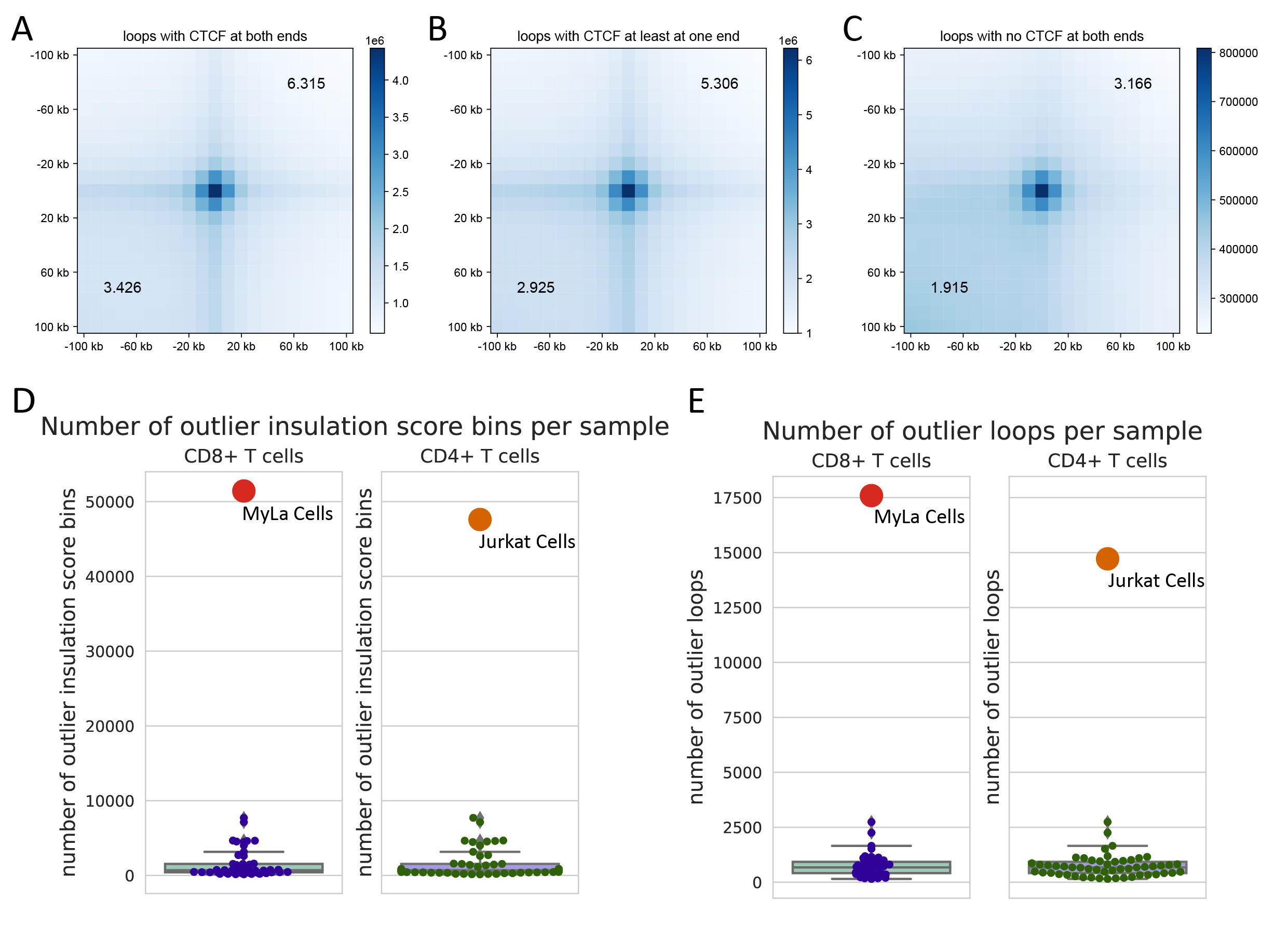

### Figure s3

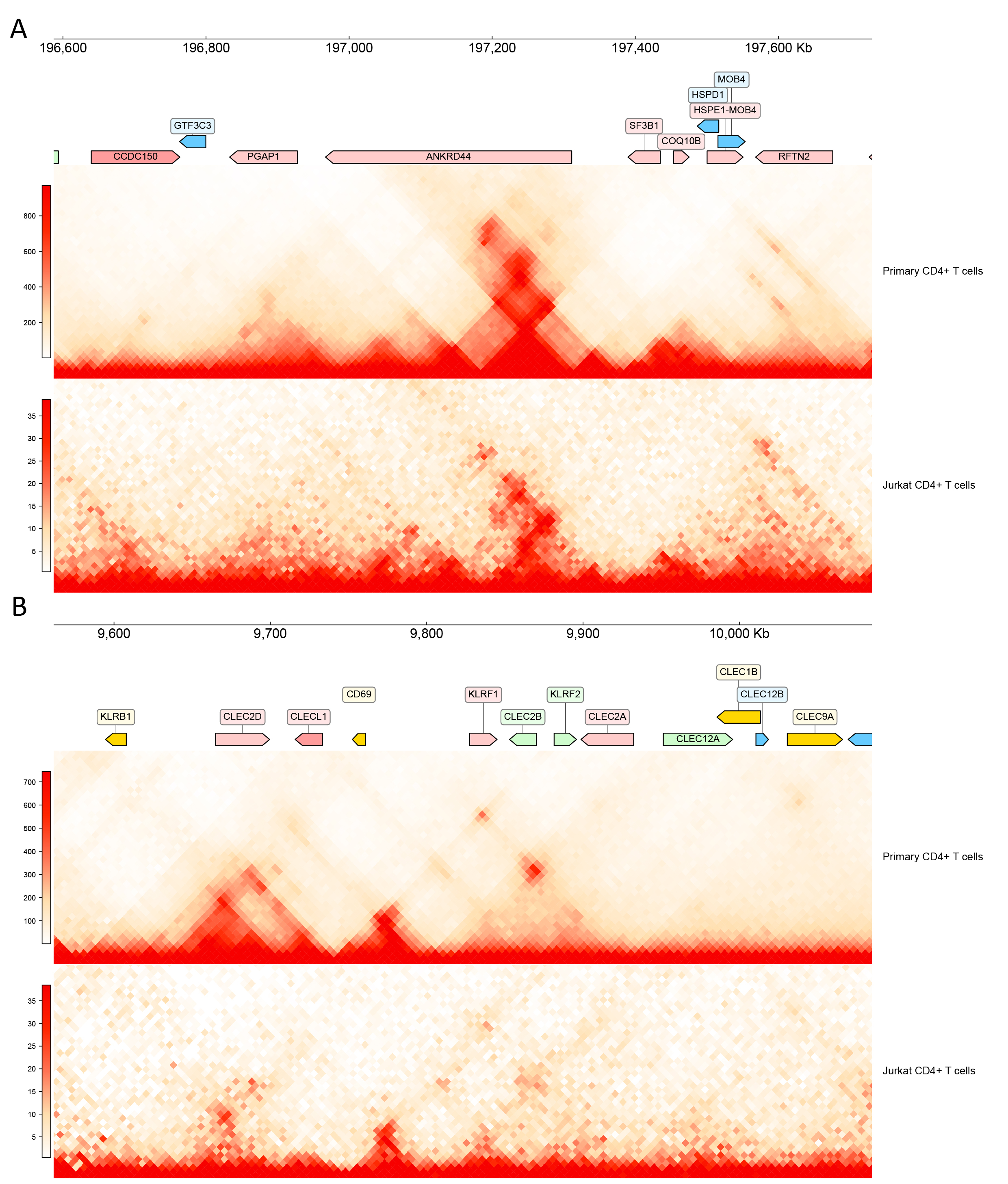

### Figure s4

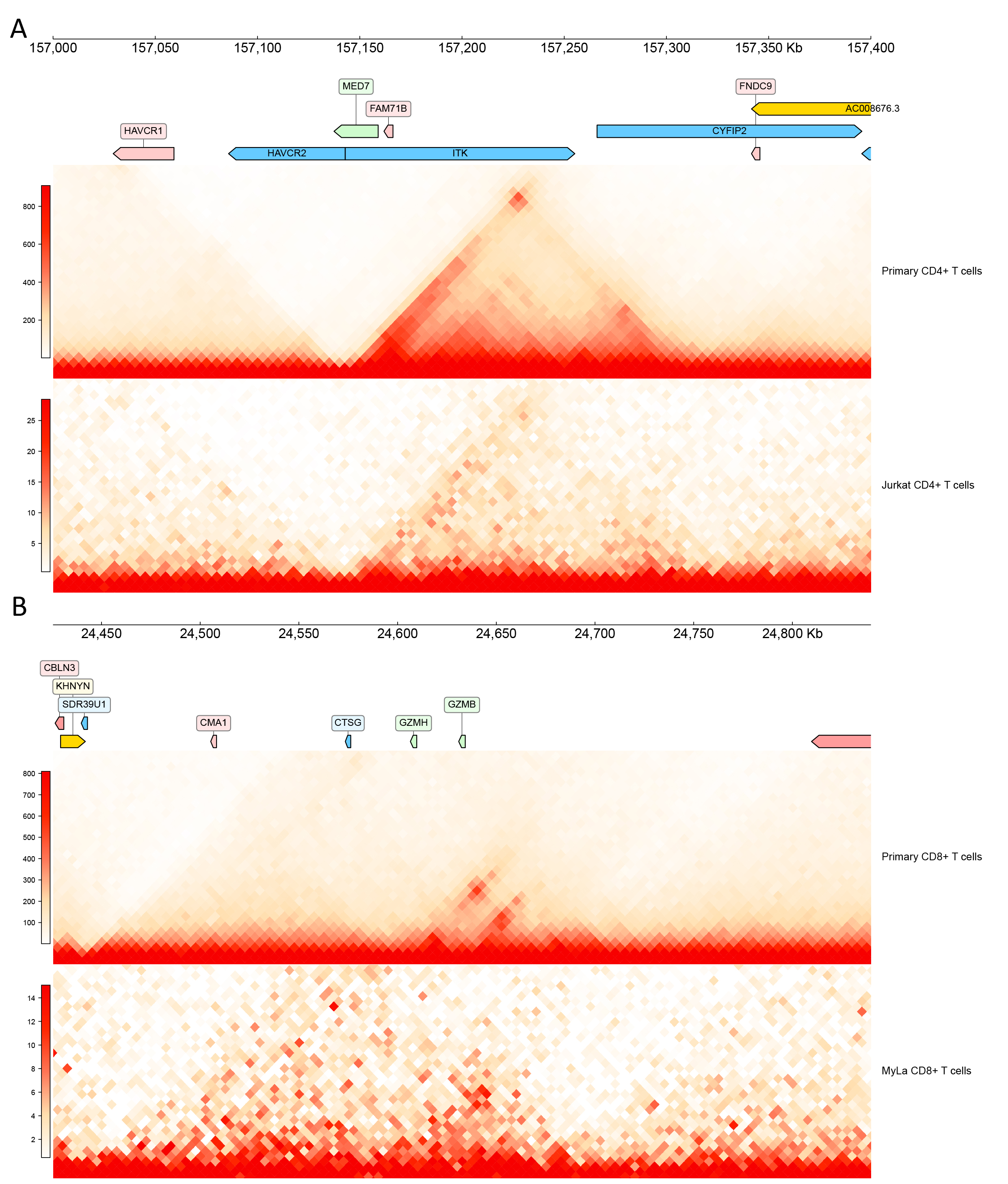

### Figure s5

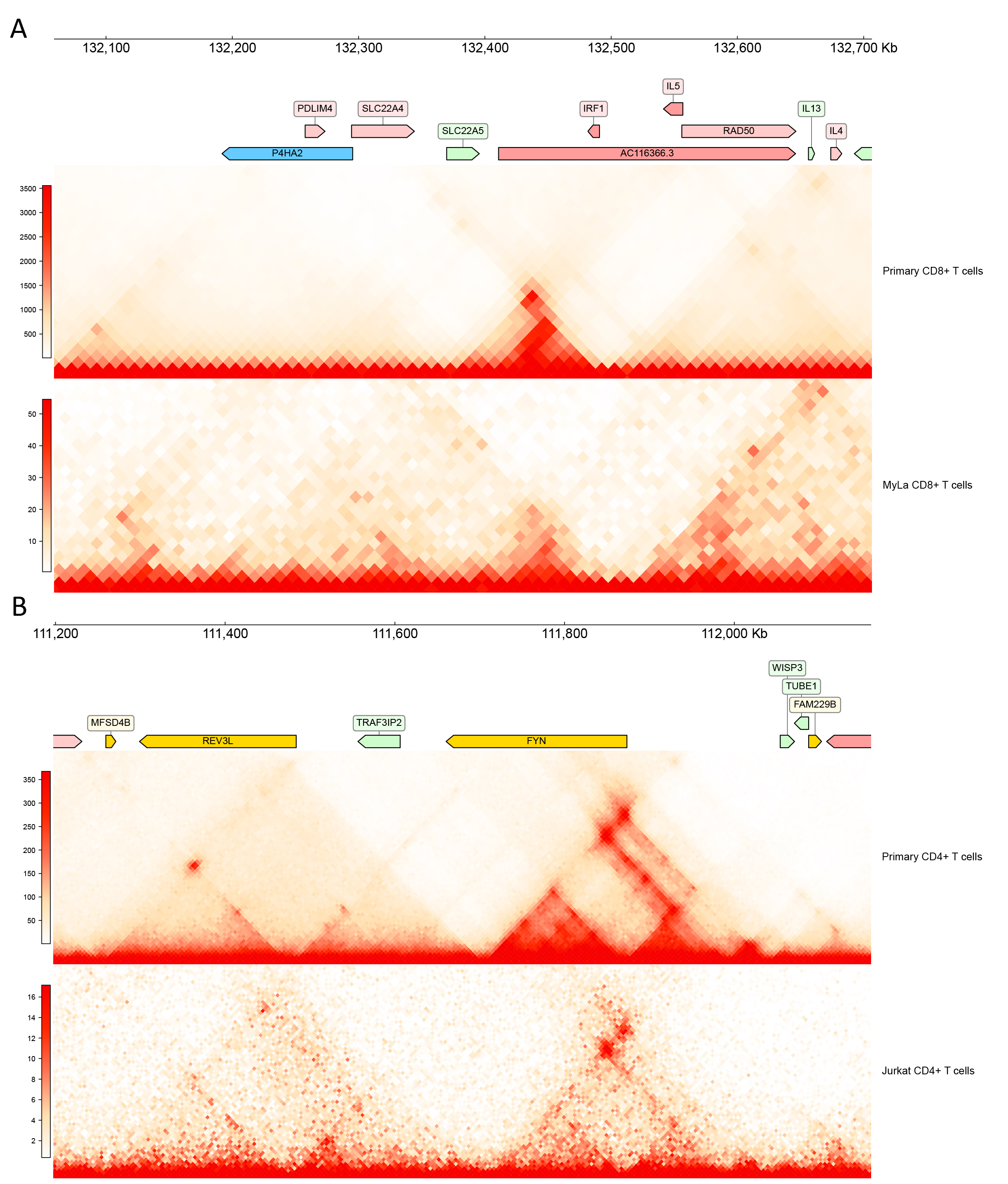

### Figure s6

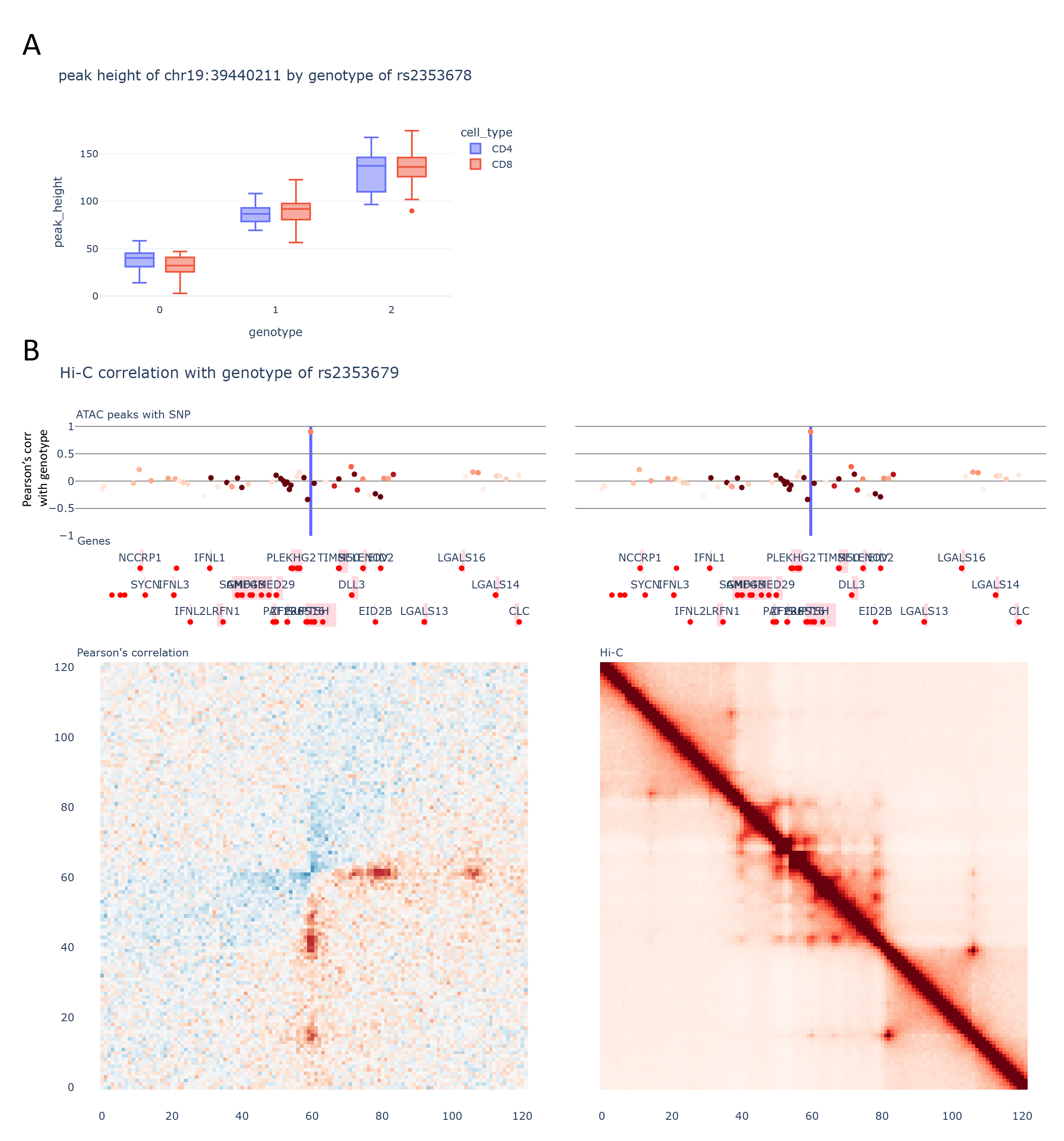

### Figure s7

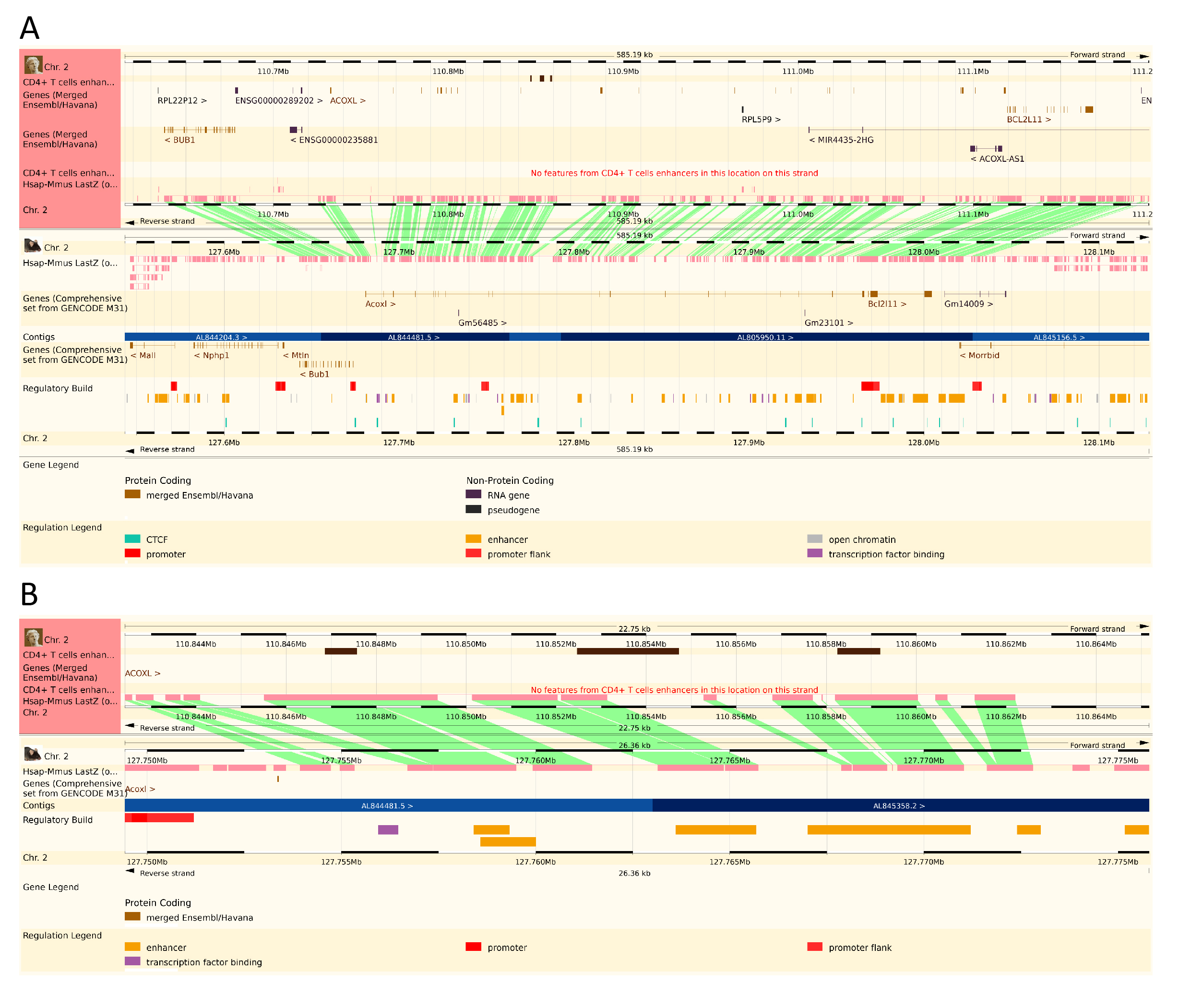
